# Covariates of success in quitting smoking in the community and secondary acute or mental health care services: a rapid systematic review

**DOI:** 10.1101/2023.01.10.23284384

**Authors:** Emma S. Hock, Matthew Franklin, Susan Baxter, Mark Clowes, Jim Chilcott, Duncan Gillespie

## Abstract

**Objectives:** To identify a comprehensive set of variables associated with quitting success among tobacco smokers contacting secondary healthcare services in the United Kingdom (UK) who are offered support to quit smoking and subsequently set a quit date.

**Design:** Rapid systematic literature review of five electronic databases.

**Setting:** Studies eligible for inclusion investigated quitting success in one of three contexts: (a) the general population in the UK; (b) people with a mental health condition; (c) quit attempts initiated within a secondary care setting.

**Interventions:** Smoking cessation intervention in a secondary care setting.

**Primary and secondary outcome measures:** Parameters from statistical analysis showing the effects of covariates on quitting success with a statistically significant (i.e., p-value <0.05) association.

**Results:** The review identified 29 relevant studies and 14 covariates of quitting success, which we grouped into four categories: demographics (age; sex; ethnicity; socio-economic conditions; relationship status, cohabitation and social network), individual health status and healthcare setting (physical health, mental health), tobacco smoking variables (current tobacco consumption, smoking history, nicotine dependence; motivation to quit; quitting history), and intervention characteristics (reduction in amount smoked prior to quitting, the nature of behavioural support, tobacco dependence treatment duration, pharmacological aids).

**Conclusions:** Fourteen data fields were identified that should be considered for inclusion in datasets and statistical analysis plans for evaluating the quitting outcomes of smoking cessation interventions initiated in secondary care contexts in the UK.

**Trial registration:** PROSPERO CRD42021254551

**STRENGTHS AND LIMITATIONS OF THIS STUDY:**

- The strengths of this review lie in the rapid but systematic approach taken and in the design of the research question and population restrictions to identify data fields important to consider in plans for the statistical analysis of the quitting outcomes of smoking cessation interventions initiated in secondary care contexts in the UK.
- The limitations lie in the compromises made as part of the rapid review approach, for example, our focus only on studies published in English, not searching grey literature, limited critical appraisal of the studies found.

## INTRODUCTION

Smoking cessation is increasingly being incorporated as a systematic and opt-out component of secondary healthcare services in the United Kingdom’s (UK’s) National Health Service (NHS), driven by a commitment to do so in the NHS’s Long Term Plan.^1–3^ The general specification of the service pathway in acute inpatient settings is: (*i*) on admission, determine if the patient smokes; (*ii*) provide advice and treatment to support patient smokers not to smoke whilst in hospital; (*iii*) provide follow-up support after discharge from hospital to support the patient to quit smoking completely. This service pathway is based on the “Ottawa Model”, following the early implementation of a hospital based tobacco dependence treatment service in Ottawa, Canada,^4^ and subsequent implementation in the UK by the CURE service in Greater Manchester.^5^ An evaluation framework for hospital based smoking cessation services in the UK was developed by consensus among UK stakeholders in acute and mental health NHS hospital Trusts,^6^ and provides a guide to the key data fields to collect for service monitoring and evaluation. However, there is no specific guidance on what data fields might be important when undertaking “deep dives” into the data to investigate factors that might influence quitting success, which in this review we generically group under the term ‘covariates’ of quitting success. Without a comprehensive list of potentially influential covariates, there is a risk that important data fields might be omitted from the routine collection of service data or from statistical analyses that aim to investigate quitting outcomes.

The current best evidence on the covariates of tobacco smoking quit success comes from a systematic review by Vangeli et al.,^7^ which examined worldwide evidence among the adult general population. The evidence presented by Vangeli et al. highlighted decreased quit success among smokers with higher nicotine dependence, smokers who smoked more cigarettes smoked each day, smokers who had made a previously unsuccessful quit attempt, and smokers who had not previously gone without smoking for a week or more. Older age and higher socio-economic status or income were also found by the review to be associated with higher quit success. However, there could also be factors specific to patient health, healthcare setting, and the features of smoking cessation interventions initiated in secondary care settings that Vangeli et al’s review of factors in the general population did not include. For example, in the British Thoracic Society’s national audits of smoking and smoking cessation intervention activities in acute NHS hospital Trusts,^8–10^ the key characteristics that were used to describe variation in whether current smokers received care for their tobacco dependence were gender, age, consultant speciality, and the patients’ route of contact with the secondary care service (elective / emergency).

This review was designed to support the evaluation of smoking cessation services in secondary care settings in the UK by identifying covariates worth considering in plans for the statistical analysis of quit success following contact with such a service. The review was based on the question: ‘What patient-, service- and setting-related factors influence the success of a quit attempt, including when initiated in a secondary care setting?’ The populations of most interest were the UK and Canada, given that the Canadian Ottawa model is the exemplar for UK services. The review question and population restrictions aimed to capture covariates of quitting success relevant to the UK general population, relevant to people with a mental health condition in any setting and in any country, and relevant to care for tobacco dependence initiated within a secondary acute or mental health service in any country. Within each study identified, the sign of the statistical coefficient for each variable investigated was taken as a measure of the direction of its association with quitting success, and the statistical significance of that coefficient at the 95% level was used to indicate if the association was potentially identified by chance or not.

## METHODS

We undertook a rapid systematic review of studies that used a statistical model to explore what covariates are associated with quitting success. We followed the rapid review approach recommended by Tricco et al.^11 12^: searching more than one database in one iteration, published literature, searches limited by date and language, research scope specified by two researchers and a health librarian, and study selection and data abstraction by one reviewer and one verifier. Quality appraisal of studies was based on whether the reporting of statistical analysis was sufficient to provide estimates of the coefficient for each variable investigated and its statistical significance at the 95% level. This rapid review approach aimed to produce a synthesis of available knowledge that was sufficient to meet the review’s aim more quickly, ensuring logistical feasibility alongside restricted timelines, while minimising risk of bias.^11 13^ The protocol is registered as PROSPERO CRD42021254551. Reporting follows PRISMA principles (http://www.prisma-statement.org/) (see the Online Supplement).

### Definition of covariates, effect size, and statistical significance

We defined a covariate of quitting success (that we term a ‘factor’) as any independent variable that can strengthen, diminish, negate, or otherwise alter the association between independent and dependent variables (in this study, the dependent variables quantify success in quitting smoking).^14^

As the dependent variable is binary (i.e., quit achieved or not by a particular time after initiating the quit attempt), we assumed that the most common statistical analysis conducted would be a form of logistic regression with effect sizes presented as odds-ratios (ORs) or unconverted beta coefficients. For descriptive purposes, when discussing effect sizes we use the following terminology whereby the binary ‘outcome’ is quitting success^15^:

- ‘Equal odds’ when OR=1; i.e., exposure does not affect odds of outcome
- ‘Higher odds’ when OR>1; i.e., exposure associated with higher odds of outcome
- ‘Lower odds’ when OR<1; i.e., exposure associated with lower odds of outcome

In keeping with the review’s aim to identify a list of potentially important covariates of quitting success, we focused on identifying which covariates have been estimated to have a statistically significant relationship with quitting success (with statistical significance defined as *p* < 0.05) rather than focussing on effect size magnitude. We define ‘no relationship’ as meaning that a covariate did not have a statistically significant relationship with quit success (i.e., *p* ≥ 0.05). We did not consider whether a relationship is causal or not, as we were interested only in association. If a study presented both univariate and multivariate analyses, we based the identification of important covariates on the multivariate analysis as this adjusts for the associations of other variables with quitting success.

### Eligibility criteria

Inclusion was restricted to studies published in peer-reviewed journals, in English, and dating from 2008, the year of the National Institute of Health and Care Excellence Guidance PH10 (for England and Wales), in which Recommendation 8 stated that smoking cessation advice and support should be available in secondary care settings for everyone who smokes. Reviews were not included, but we checked references for any relevant studies. We included studies that presented statistical estimates of the effects of covariates on the success of a quit attempt.

We searched for studies statistically assessing quit attempts in three contexts: (a) the general population instigated in any setting within the UK; (b) people with a mental health condition instigated in any setting and in any country; (c) initiated within a secondary acute or mental health service in any country. The scope of (a) was limited to the UK for relevance and feasibility given the large number of studies worldwide.

### Information sources

Searches were conducted in April 2021. A focused search strategy combining free-text terms with subject headings (e.g., MeSH) was run and translated for optimal effectiveness across the following databases: MEDLINE (including In-Process and Epub ahead of print); EMBASE; PsycINFO (all via Ovid); CINAHL (via EBSCO) and the Cochrane Library.

### Search process

The search strategy was constructed around the facets of: Smoking cessation AND quitting success AND (UK OR mental health OR hospital setting). Due to the time-constrained nature of this review, searches prioritised specificity over sensitivity, but to mitigate the risk of missing relevant papers the strategy was validated against six studies already known by the authors to be potentially relevant: Le Grande et al.,^16^ Lubitz et al.,^17^ Ussher et al.,^18^ Smit et al.,^19^ Vangeli et al.,^7^ and Zhou et al.^20^ All six studies were retrieved by the search (see the Online Supplement). Database search results were extracted directly to reference management software.

### Study selection

Screening for studies relevant to each of our three contexts (a–c) was performed simultaneously, with included studies marked for relevance to each. Titles and abstracts were screened by one of three reviewers (EH, MF or SB); 70% of abstracts were checked by another reviewer (EH or MF).

Full texts were assessed for inclusion by one reviewer and checked by another reviewer (EH or MF). Disagreements were resolved through discussion, with no need to involve a third reviewer.

### Data extraction and synthesis

EH and MF designed and tested a spreadsheet for data extraction. Data were extracted and charted by EH and checked in regular meetings with MF and DG. The following data items were extracted: Reference information (first author and date), study type, country, setting (e.g., hospital type/department/ward), participant baseline characteristics (e.g., age, sex, socio-economic status, reason for admission, cigarettes/day smoked, number of previous quit attempts, nicotine dependence), measure of quit success (point prevalence abstinence or continuous abstinence, any time point but recorded separately per time-point). Relevant characteristics of the analysis were noted. For example, method of data collection, sample size, time horizon, cessation time-point, measure of abstinence, whether ORs and model coefficients were captured, the model type, and whether a univariate or multivariate model. Detailed statistical results were also extracted: the whole model, where reported, including intercept and other coefficients, dependent and independent variable, any reported *p*-values, and goodness of fit statistics, if reported.

During the data extraction process, we began to develop an organisational framework by categorising studies according to our three contexts, the covariates investigated and their effects on quit success. The organisational framework was then revised as results synthesis progressed. Covariates were grouped according to our final organisational framework.

## RESULTS

From 2,499 retrieved records, 29 studies were included in the synthesis (**Error! Reference source not found**.), representing 21 studies relating to the UK general population context, six studies relating to mental health in the UK or Canada, and two studies relating to secondary care in the UK or Canada. A list of excluded studies with reasons is available in Supplementary Table S1.

### Description of included studies

The characteristics of the included studies are summarised in Supplementary **Error! Reference source not found**.; Supplementary Table S3 shows study participants characteristics. Most studies had prospective, cross-sectional or retrospective designs; three studies were randomised controlled trials (RCTs).

### Methodological differences between studies

Methodological differences are reported in Supplementary Table S4. Smoking cessation was assessed in a variety of different ways across studies. The time horizon for reporting smoking abstinence following a quit attempt ranged from 2 weeks to 1 year. Abstinence was assessed as both point-prevalent and continuous, both by self-report (most frequently used for continuous abstinence) and validated by expired air carbon monoxide (CO; most frequently used to verify 7-day or 2-week point-prevalent abstinence, at ≤10 or ≤8 ppm). If a study conducted separate analyses for different durations of abstinence following a quit attempt, we reported the findings from each analysis independently. All studies reported odds-ratios from a logistic regression, and two studies reported beta coefficients.

In terms of sample, the majority of UK studies were of the general population (15 studies) or community smoking cessation services (four studies), with three studies examining samples with specific characteristics (i.e., pregnant women, people aged 25–59 years, and English residents of Bangladeshi origin; see Supplementary Table S3). Mental health population studies were from Canada and sampled from people attending community mental health services (four studies) or from the general population (two studies). The two secondary care studies recruited participants from a Canadian hospital-based smoking cessation clinic or UK cardiac rehabilitation setting data.

### Covariates of success in quitting tobacco smoking

Figure 2 summarises the covariates that had a statistically significant relationship with quit attempt success. Supplementary Table S5 summarises the relationships between covariates and quit success. Supplementary Table S6 provides a full description of the size and direction of covariate effects and the corresponding statistical significance.

**Figure 1.**
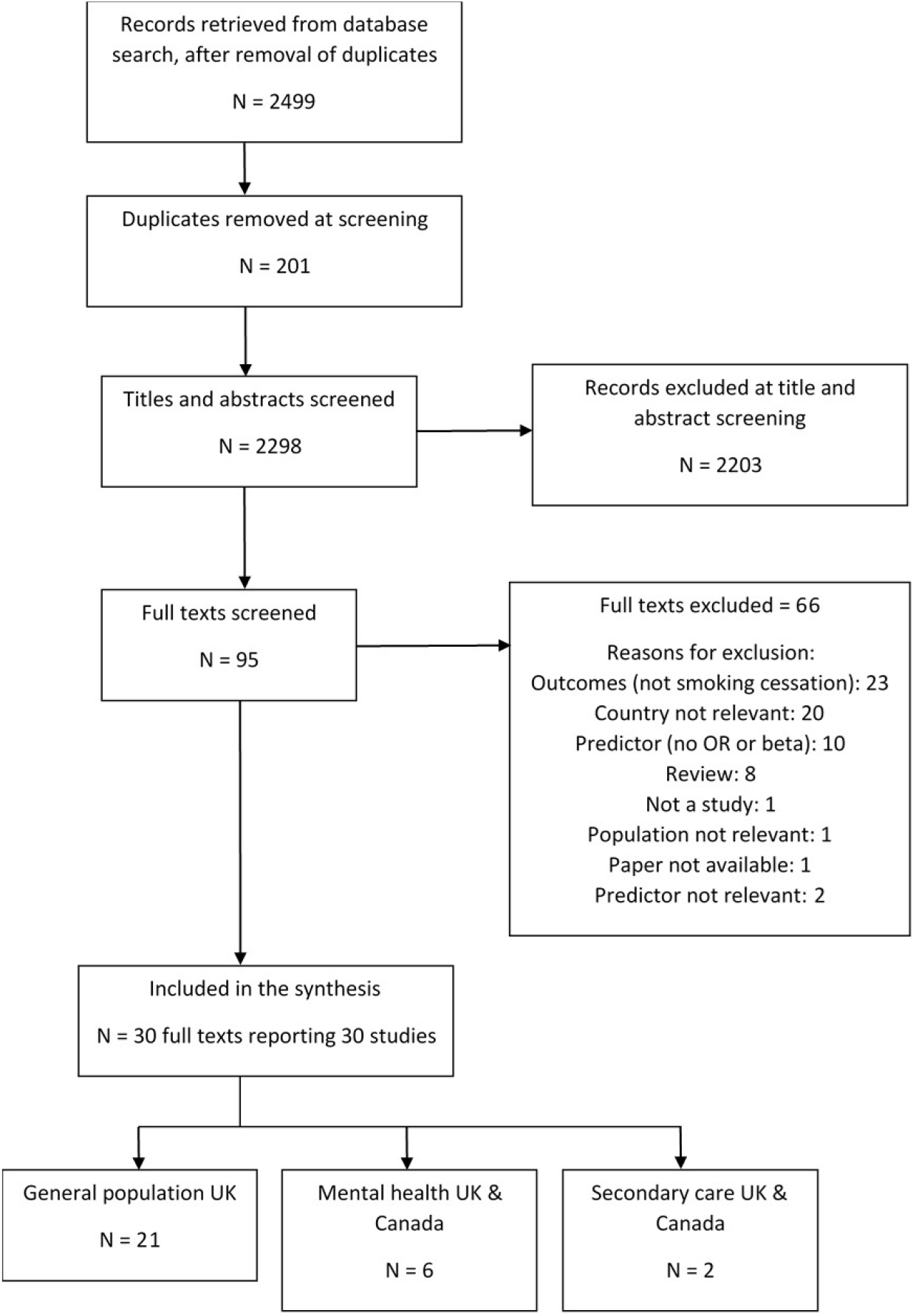
PRISMA flow diagram of study inclusion.

**Figure 2.**
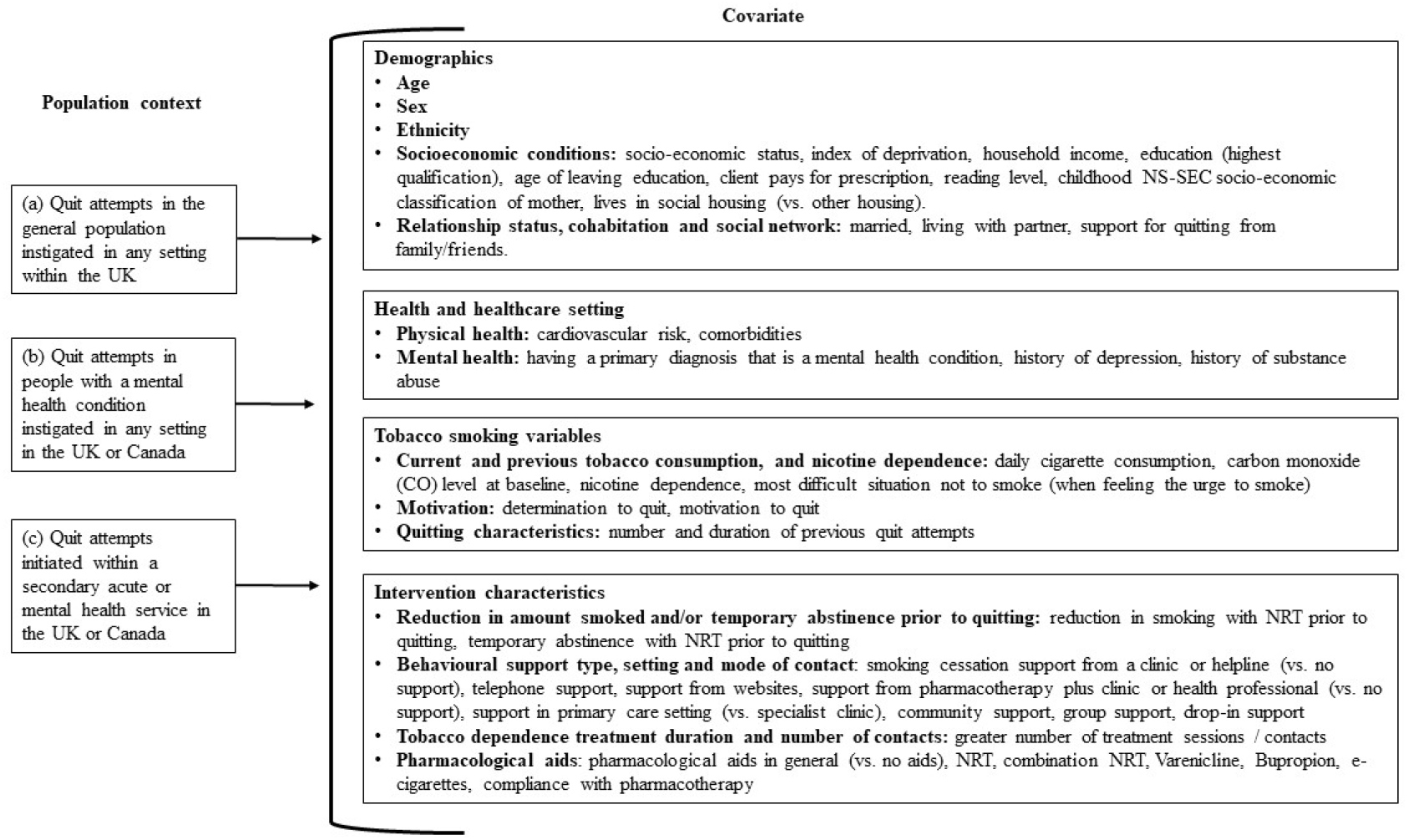
List of covariates found to have a statistically significant association with quitting success in at least one study. Supplementary Table S6 provides a full description of the size and direction of covariate effects and the corresponding statistical significance.

#### Demographics

Overall, 16 studies included demographic covariates; the factors related to quit outcome were age, sex, ethnicity, socioeconomic conditions, smoker’s relationship status, cohabitation and social network situation (Supplementary Table S5; Supplementary Table S6).

##### Age

All studies showed higher odds (i.e., of quit success) with increasing age.^21–25^ Six analyses reported in five papers found no relationship between age and quit success in the UK general population, ^26–30^ two studies found no relationship for age in people with mental health conditions,^31 32^ and two studies found no relationship in a secondary care setting.^33 34^

##### Sex

There were inconsistent findings for sex: in the UK general population, three studies reported higher odds of quitting success for males^21 23 28^ and two studies reported higher odds of quitting success for females.^24 26^ Two studies in an outpatient setting (cardiology and mental health services) found higher odds of quitting success in males.^33 35^ Six studies found no relationship between sex and quitting success in the UK general population,^22 25 27 29 30 36^ and two studies found no relationship in people with mental health conditions.^31 32^

##### Ethnicity

One study reported higher odds of quitting success for Black ethnicity vs. White British ethnicity.^23^ One study reported no relationship between ethnicity and quitting success in the UK general population.^29^

##### Socioeconomic characteristics

There was a varied definition of socioeconomic characteristics in the studies identified. Higher odds of quitting success were reported for people: with higher social grades^23 27–29 37^; living in less deprived areas^25^; have higher income^36 38^; higher occupational grades^21 38^; more education^26 38^; who paid for prescriptions vs. were exempt^21 22^; had a higher reading level^29^; people whose mothers worked in higher grade occupations during their childhood^38^; and people who did not live in social housing^39^. In the UK general population, one study reported no relationship between quitting success and the geographic Index of Multiple Deprivation (IMD) score for the location of the smoking cessation service,^21^ five studies reported no relationship between quitting success and education,^24 26 29 36 38^ one study for prescription exemption status,^24^ and one study for employment status.^24^ In a secondary care setting, two studies reported no relationship between quitting success and the employment status of patients.^33 34^

##### Relationship status, cohabitation and social network

A study in the UK general population found higher odds of quitting success for people who were single, divorced or separated vs. were married or living with a partner^24^. However, a study of patients in care for cardiac rehabilitation found higher odds of quitting success for people who were married vs. single^34^. In the UK general population, studies reported finding no relationship between quitting success and marital status,^29^ cohabitation status,^38^ or number of household smokers.^23 24^ One study of people with severe and persistent mental illness reported higher odds of quitting success for people with more social support for quitting from family/friends.^31^

#### Health and healthcare setting

There were eight studies that investigated the association between quitting success and the smoker’s health or the healthcare setting in which the quit attempt was instigated; five reported covariates that had statistically significant relationships to quitting success (Supplementary Table S5; Supplementary Table S6): level of cardiovascular risk; number of comorbidities; having a mental health diagnosis; having a history of depression; having a history of substance abuse.

##### Physical health

One study in an outpatient setting reported higher odds of quitting success for patients with low (vs. moderate or high) cardiovascular risk and patients with fewer comorbidities.^34^ However, no relationship was found between quitting success and moderate (vs. high) cardiovascular risk.^34^ Another study found no relationship between quitting success and the number of comorbidities that a patient had.^33^ One study reported no relationship between the clinical setting in which the patient was located at the time that they were referred to stop smoking support (Cardiology services/clinics vs. Respirology services/clinics vs. other hospital services/clinics).^33^

##### Mental health

Lower odds of quitting success were reported for people with: a primary diagnosis of anxiety disorder vs. no disorder^32^; recurrent, current or recent depression vs. no history of depression^40^; history of opiate abuse vs. history of alcohol abuse^32^; history of alcohol abuse, opiate abuse and marijuana abuse vs. no history of substance abuse.^41^ No relationship with quitting success was reported in three studies that investigated primary mental health diagnosis,^31 35 41^ two studies of PHQ-9 score,^31 42^ one study of having a history of substance abuse,^31^ one study of HADS anxiety score and HADS depression score,^34^ and one study of history of psychiatric disorder and history of co-occurring substance use and psychiatric disorder.^33^

#### Tobacco smoking variables

There were 17 studies in this category; 14 reported factors significantly related to quitting success (Supplementary Table S5; Supplementary Table S6): daily cigarette consumption, carbon monoxide (CO) level at baseline, level of nicotine dependence, the most difficult situation not to smoke, determination / motivation to quit, and the history of previous attempts to quit smoking.

##### Current and previous cigarette consumption

Higher odds of pregnant women quitting smoking successfully were reported among women with lower pre-pregnancy cigarette consumption.^38^ No relationship between quitting success and the daily cigarette consumption prior to quitting was identified in one study in the UK general population,^28^ two studies of people with a mental health condition^31 32^ and one study in a secondary care setting.^33^ No relationship between quitting success and the age at which someone started to smoke regularly (age at smoking initiation) was reported by one study in the UK general population,^29^ two studies in people with a mental health condition,^31 32^ and one study in a secondary care setting.^33^

##### Carbon monoxide (CO) level

The single study to find a relationship between quitting success and CO level prior to quitting was of a tailored smoking cessation programme for individuals with substance use disorders and mental illness; lower CO levels at when the quit attempt began had higher odds of quitting success.^41^ No relationship between quitting success and CO level was found by one study in people with a mental health condition,^32^ and one study in a secondary care setting.^33^

##### Level of nicotine dependence

The 11 studies which identified statistically significant associations between quitting success and nicotine dependence prior to the quit attempt found mixed results: higher odds of quitting in smokers with lower nicotine dependence was found by nine studies in the UK general population^21 24–29 36 43^ and two studies of smoking cessation delivered in an outpatient setting.^31 32^ No relationship between quitting success and nicotine dependence was found by one study in the UK general population,^23^ two studies in people with a mental health condition,^35 41^ and one study in a secondary care setting.^33^ One study in the UK general population found higher odds of quitting success in smokers whose most difficult situation not to smoke was when feeling the urge to smoke, but the same study found no relationship with quitting success for when socialising, first thing in the morning, when angry or frustrated, when relaxing, and for ‘any other reason’.^29^ One study found no relationship between quitting success and the reported enjoyment of smoking.^27^

##### Motivation to quit

Two studies in the UK general population found higher odds of quitting successfully for smokers who reported a determination to quit^23^ or being motivated to quit.^36^ No relationships between quitting success and reported readiness to quit were found in one study in the UK general population,^29^ one study in people with a mental health condition,^31^ and one study in a secondary care setting.^33^ One UK general population study found no relationship between quitting success and the reported reasons for quitting, main advantage of quitting, or main disadvantage of quitting.^29^

##### Quitting characteristics

In terms of previous quit attempts, three studies in the UK general population^26 28 29^ and one study in a mental health setting^32^ found higher odds of quitting successfully among smokers who had made more previous quit attempts or had previously been abstinent for longer periods. Specifically, higher odds of quitting successfully were found among those who had previously quit smoking for 3 months or more,^29^ made ≥2 quit attempts in the past 6 months,^28^ and had a longer duration of abstinence at the last attempt to quit.^26 32^ Three studies in the UK general population reported no relationship between quitting success and the number or duration of previous quit attempts,^24 28 44^ as did one study in people with a mental health condition,^31^ and one study in an outpatient setting.^33^ One study in a UK general population reported no relationship between success in the current quit attempt and the time since the start of the last unsuccessful quit attempt.^28^

#### Intervention characteristics

There were 21 studies that investigated the influence on quitting success of characteristics of the attempt to quit smoking; 17 studies reported factors significantly related to the success of quit attempts (Supplementary Table S5; Supplementary Table S6). Factors related to the behaviour and choices of the individual smokers were whether smokers reduced or temporarily abstained from smoking before making a quit attempt, and various descriptors of the nature of support for the quit attempt. Pharmacological characteristics of the quit attempt were the type of pharmacological aid use, whether this was used alongside behavioural support, and the degree of compliance of the smoker making the quit attempt with the recommended guidelines for use of the pharmacotherapy chosen.

##### Reduction in amount smoked and/or temporary abstinence before quitting

Two studies found higher odds of quitting successfully for smokers who reduced the amount they smoked before attempting to quit smoking,^28 45^ including if this was with the support of pharmacotherapy.^45^ One study found no relationship between quitting success and whether the quit attempt was spontaneous, i.e. initiated as soon as the decision to quit has been made (compared with not making a spontaneous quit attempt),^28^ and one study found no relationship between quitting success and whether the smoker reduced the amount smoked prior to quitting (compared with quitting without first reducing the amount smoked).^26^

##### Behavioural support type, setting and mode of contact

For the UK general population, higher odds of quitting were found for smokers who used a smoking cessation clinic and websites (compared with no support),^39 46^ for smokers who used pharmacotherapy alongside help from a health professional or specialist smoking cessation advisor (compared with no support),^46^ and for smokers who received support in specialist clinics,^21 44^ in the community (compared with other settings),^24 25^ and with group support (compared with one-to-one or other support).^21 22^ Lower odds of quitting were reported for smokers who used drop-in support (compared with one-to-one support),^44^ and telephone support (compared with no support).^39^ Other studies found no relationships between quitting success and the receipt of in-person behavioural support,^39^ the use of self-help materials,^39^ having one-to-one support,^47^ the setting of support for smoking cessation,^21 22 25 44^ having group therapy, or receiving support from a doctor or other health professional.^46^

##### Tobacco dependence treatment duration and number of contacts

Higher odds of quitting success were associated with the number of contacts that a smoker had with a stop smoking advisor in the UK general population,^23^ and in studies of people with a mental health condition.^32 35 41^ Other studies found no relationship between quitting success and treatment duration or number of contacts.^21 31 33^

##### Pharmacological aids

In the UK general population, higher odds of quitting success were found for smokers who used NRT (compared with no NRT/no cessation aids),^21 39 44^ combination NRT (compared with single NRT),^30^ varenicline (compared with no varenicline, no medication, or NRT),^21 25 39 44^ bupropion (compared with no medication and NRT),^21 24^ and for the use of any pharmacotherapy in general.^46 48^ There were also higher odds of quitting success with the use of e-cigarettes (compared with no e-cigarettes, no cessation aid, and NRT).^36 39 47^ There was also evidence in the UK general population of higher odds of quitting successfully when smokers have greater compliance with the recommended guidelines for pharmacotherapy use.^23^ One study in the UK general population found lower odds of quitting successfully for smokers who bought NRT over the counter (compared with no cessation aids).^48^ Other studies in the UK general population found no relationships between quitting success and the use of prescription NRT,^39^ NRT bought over the counter,^39^ bupropion,^39 44^ or e-cigarette use.^36^ For people with a mental health condition, no relationship with quitting success was found for the use of pharmacotherapy,^31 32 35^ or the number of weeks of NRT, varenicline and bupropion use.^32^

## DISCUSSION

The review has identified a list of covariates worth considering in plans for the statistical analysis of quitting success following a smoking cessation intervention initiated in a secondary care setting in the UK. The findings support and supplement the previous reviews that have investigated covariates of quitting success, and add to the evaluation framework for hospital based smoking cessation services in the UK^6^ by highlighting the data fields important to consider in “deep dives” into service data to investigate the reasons for variation in quitting outcomes.

### Strength and limitations

The strengths of this review lie in the rapid but systematic approach taken^11 12^ and in the design of the research question and population restrictions to be specific to smoking cessation interventions initiated in a secondary care setting in the UK. The limitations lie in the compromises made as part of the rapid review approach, for example, our focus only on studies published in English, not searching grey literature, limited critical appraisal of the studies found. the review only included studies from the UK and Canada, which was intended to limit the influence of variation in service delivery internationally, while noting our interest was specific to the UK. Whilst this restriction increased relevance, only two studies were identified from a secondary healthcare setting. It is possible that expanding the search worldwide would have identified more covariates specific to understanding the influence of health and the healthcare setting on quitting success. However, healthcare systems differ widely worldwide, and our decisions to limit the scope of this review are in line with recommended best practice for rapid reviews.^11 12^

### Informing real-world data collection: supporting clinical care and public health policy

Improvement of smoking cessation interventions embedded into NHS secondary care services requires the use of real-world data for service monitoring and ongoing evaluation. There will be incremental improvement in services over time, including attempts to address factors observed to influence the success of quit attempts. This review provides a starting point for understanding what data fields might be important to collect to ensure that sufficient information is available to guide activities aimed at service improvement. The NICE real-world evidence framework^49^ encourages service evaluators to identify the data fields needed through a systematic, transparent and reproducible search. The current review of the covariates of quitting success is part of that systematic approach and could aid the planning of data fields to be collected.

### Evidence-based care: trial-based and real-world evidence

When conducting an evaluation of intervention efficacy or comparative effectiveness, be it based on a randomised or non-randomised study design (noting service evaluations are not permitted to randomise patients to treatment assignment), developing a statistical analysis plan is an important step towards reducing potential bias in the evidence base.^49^ Service evaluations and associated real-world evidence are often dependent on the real-world data available, hence the importance of considering which covariates to collect data on. For a statistical analysis plan, the interest is usually in adjusting estimates of service outcomes for the influence of confounding variables, but investigations can become more complex by situating covariates within a causal framework for evaluating service outcomes, for example using directed acyclic graphs.^49^ The list of covariates identified in the current review could aid the development of a range of plans for statistical analysis to inform the evidence-base, focussed either on association or causality depending on the intention of the analysis and required evidence-base.

### Understanding service complexity: informing adaptive logic models

There is increasing recognition in real world implementation and evaluation of healthcare interventions of the complexity of even seemingly “simple” treatments. Healthcare has been described as a complex adaptive system which requires understanding of multiple elements and the way in which they interact, in order to lead to transformation.^50^ In common with many evaluations, evaluations of tobacco dependence treatment services in the UK draw on a theory of change approach in order to aid understanding of implementation and the effects of the tobacco dependence treatment service on outcomes for smoking and health.^51^ The data fields identified during this review help to inform the development of service logic models,^52^ which act as a visual summary of the complexity by which the intervention produces outcomes. These models can help to build our conceptualization and understanding of hypothesized causal links underpinning quitting smoking.^53^

## CONCLUSION

Fourteen broad categories of covariate were identified as having a statistically significant association with the success in quitting smoking and therefore worth considering in plans for the statistical analysis of quit success following contact with a smoking cessation intervention initiated within secondary healthcare services in the UK. These covariates also indicate the data fields it might be important to collect as part of the ongoing monitoring and evaluation of such services.

## Supporting information

PRISMA abstract checklist

PRISMA checklist

## Data Availability

The spreadsheet containing data extracted from included studies is available on request.

Supplementary Information: PRISMA checklist & PRISMA abstract checklist

Supplement 1 – search strategies in full

Supplement 2 – results tables

- Supplementary Table S1: Studies excluded at full text screening
- Supplementary Table S2. Characteristics of included studies.
- Supplementary Table S3: Participant baseline characteristics of included studies
- Supplementary Table S4: Outcome measurement and analyses in included studies
- Supplementary Table S5. Relationships between covariates and quit success.
- Supplementary Table S6: Covariates of quitting outcomes – full results summary

## ACKNOWLEDGEMENTS

The review was initiated as part of a Yorkshire Cancer Research commissioned service evaluation of the QUIT hospital-based tobacco dependence treatment service (https://sybics-quit.co.uk/). The authors would like to thank the Programme Director of the QUIT service, Lisa Wilkins, and the teams implementing the service for their support with the QUIT service evaluation. The authors thank Debbie Robson for support in developing the review and John Holmes for comments to improve the manuscript.

For the purpose of open access, the author has applied a CC BY public copyright licence to any Author Accepted Manuscript version arising from this submission.

## Data availability statement

The spreadsheet containing data extracted from included studies is available on request.

## Author contributions

- Emma S. Hock: Writing – Original Draft Preparation (equal), Writing – Review & Editing (lead), Investigation - evidence searches and collation (lead)
- Matthew Franklin: Funding Acquisition (supporting), Conceptualization (lead), Writing – Review & Editing (supporting), Investigation - evidence searches and collation (supporting), Validation (equal), Project Administration (supporting), Supervision (equal)
- Susan Baxter: Funding Acquisition (supporting), Writing – Review & Editing (supporting), Investigation - evidence searches and collation (supporting)
- Mark Clowes: Methodology - designing the searches (lead)
- Jim Chilcott: Funding Acquisition (supporting), Writing – Review & Editing (supporting), Validation (equal)
- Duncan Gillespie: Funding Acquisition (lead), Writing – Original Draft Preparation (equal), Writing – Review & Editing (lead), Project Administration (lead), Supervision (equal)

## ETHICS APPROVAL

Ethics approval was not required for this review study.

