## Supplement 1 for "Covariates of success in quitting smoking in the community and secondary acute or mental health care services: a rapid systematic review"

**Online supplement – Search strategies**

**MEDLINE** including In-Process and Epub ahead of print (via Ovid, 19^th^ April 2021)

| 1. | *smoking cessation/ or *"tobacco use cessation"/ or ((quit* or stop* or cessation) adj2 smok*).ti. |
| --- | --- |
| 2. | (smok* and (brief adj2 (advi* or intervention*))).ti,hw,kw. |
| 3. | 1 or 2 |
| 4. | ((predict* or factor* or correlate*) adj3 (success* or lapse* or fail* or outcome* or quit* or cessation)).ti,ab. |
| 5. | 3 and 4 |
| 6. | limit 5 to (english language and yr="2008 -Current") |
| 7. | exp Great Britain/ |
| 8. | (national health service* or nhs*).ti,ab,in. |
| 9. | (english not ((published or publication* or translat* or written or language* or speak* or literature or citation*) adj5 english)).ti,ab. |
| 10. | (gb or "g.b." or britain* or (british* not "british columbia") or uk or "u.k." or united kingdom* or (england* not "new england")  or northern ireland* or northern irish* or scotland* or scottish* or ((wales or "south wales") not "new south wales") or  welsh*).ti,ab,jw,in. |
| 11. | 7 or 8 or 9 or 10 |
| 12. | (exp africa/ or exp americas/ or exp antarctic regions/ or exp arctic regions/ or exp asia/ or expoceania/)  not (exp great britain/ or europe/) |
| 13. | 11 not 12 |
| 14. | 6 and 13 |

**EMBASE** (via Ovid, 20^th^ April 2021)

| 1. | *smoking cessation/ or *"tobacco use cessation"/ or ((quit* or stop* or cessation) adj2 smok*).ti. |
| --- | --- |
| 2. | (smok* and (brief adj2 (advi* or intervention*))).ti,hw,kw. |
| 3. | 1 or 2 |
| 4. | ((predict* or factor* or correlate*) adj3 (success* or lapse* or fail* or outcome* or quit* or cessation)).ti,ab. |
| 5. | 3 and 4 |
| 6. | exp Great Britain/ |
| 7. | (national health service* or nhs*).ti,ab,in. |
| 8. | (english not ((published or publication* or translat* or written or language* or speak* or literature or citation*) adj5  english)).ti,ab. |
| 9. | (gb or "g.b." or britain* or (british* not "british columbia") or uk or "u.k." or united kingdom* or (england* not "new england")  or northern ireland* or northern irish* or scotland* or scottish* or ((wales or "south wales") not "new south wales")  or welsh*).ti,ab,jw,in. |
| 10. | 6 or 7 or 8 or 9 |
| 11. | (exp africa/ or exp americas/ or exp antarctic regions/ or exp arctic regions/ or exp asia/ or expoceania/)  not (exp great britain/ or europe/) |
| 12. | 10 not 11 |
| 13. | exp Mental Disorders/ |
| 14. | (mental health or mental* ill* or mental disorder* or psychot* or psychos* or psychiatr* or schizophren* or bipolar or  bi-polar or depression or anxiety).mp. |
| 15. | exp Hospitals/ |
| 16. | (hospital* or ward* or acute setting*).mp. |
| 17. | 12 or 13 or 14 or 15 or 16 |
| 18. | 5 and 17 |
| 19. | limit 18 to (english language and yr="2008 -Current") |
| 20. | remove duplicates from 19 |
| 21. | limit 20 to english language |
| 22. | limit 21 to embase |

**PsycINFO** (via Ovid, 20^th^ April 2021)

| 1. | ((quit* or stop* or cessation) adj2 smok*).ti,hw. |
| --- | --- |
| 2. | (smok* and (brief adj2 (advi* or intervention*))).ti,hw. |
| 3. | 1 or 2 |
| 4. | ((predict* or factor* or correlate*) adj3 (success* or lapse* or fail* or outcome* or quit* or cessation)).ti,ab. |
| 5. | 3 and 4 |
| 6. | (UK or united kingdom or great britain or england or scotland or northern ireland or wales or national health  service or NHS).af. |
| 7. | (mental health or mental* ill* or mental disorder* or psychot* or psychos* or psychiatr* or schizophren* or  bipolar or bi-polar  or depression or anxiety).mp. |
| 8. | (hospital* or ward* or acute setting*).mp. |
| 9. | limit 5 to (english language and yr="2008 -Current") |
| 10. | 6 or 7 or 8 |
| 11. | 9 and 10 |

**Cochrane Library**, searched 20^th^ April 2021

Search Name:	YCR smoking

Date Run:	20/09/2022 10:38:35

Comment:

ID	Search	Hits

#1	MeSH descriptor: [Smoking Cessation] explode all trees

#2	(((quit* or stop* or cessation) near/2 smok*)):ti OR (((quit* or stop* or cessation) near/2 smok*)):kw (Word variations have been searched)

#3	((smok* and (brief near/2 (advi* or intervention*)))):ti,ab,kw (Word variations have been searched)

#4	(((predict* or factor* or correlate*) near/3 (success* or lapse* or fail* or outcome* or quit* or cessation))):ti,ab,kw (Word variations have been searched)

#5	#1 or #2 or #3

#6	MeSH descriptor: [United Kingdom] explode all trees

#7	(UK or united kingdom or brit* or england or scotland or wales or northern ireland):ti,ab,kw (Word variations have been searched)

#8	MeSH descriptor: [Mental Disorders] explode all trees

#9	(UK or united kingdom or brit* or england or scotland or wales or northern ireland):ti,ab,kw (Word variations have been searched)

#10	MeSH descriptor: [Hospitalization] explode all trees

#11	(hospital* or ward* or acute setting*):ti,ab,kw (Word variations have been searched)

#12	#6 or #7 or #8 or #9 or #10 or #11

#13	#4 and #5 and #12

| **CINAHL** (via EBSCO, 20^th^ April 2021) |  |  |
| --- | --- | --- |
| **#** | **Query** | **Limiters/Expanders** |
| S7 | S5 AND S6 | Expanders - Apply equivalent subjects Search modes - Boolean/Phrase |
| S6 | ( brit* or uk or "united kingdom" or england or scotland or wales or northern ireland ) OR ( hospital or acute setting or inpatient* or ward ) OR ( mental health or mental illness or mental disorder or psychiatric illness or schizophrenia or bipolar or bi-polar or anxiety or depression ) | Limiters - Published Date: 20080101-20211231 Expanders - Apply equivalent subjects Search modes - Boolean/Phrase |
| S5 | S1 AND S2 |  |
| S4 | S1 AND S2 |  |
| S3 | S1 AND S2 |  |
| S2 | ( success* or lapse* or fail* or outcome* or quit* or cessation ) AND ( predict* or factor* or correlate* ) |  |
| S1 | ( (MM "Smoking Cessation") OR (MM "Smoking Cessation Programs") OR (MM "Tobacco Use Cessation Products+") OR (MM "Smoking Cessation Assistance (Iowa NIC)") ) OR TI ( ((quit* or stop* or cessation) near/2 smok*) ) OR ( (smok* and (brief near/2 (advi* or intervention*))) ) |  |
