## Supplement 2 for "Covariates of success in quitting smoking in the community and secondary acute or mental health care services: a rapid systematic review"

**Online supplement – Results tables**

### Supplementary Table S1: Studies excluded at full text screening

| **Paper** | **Reason for exclusion** |
| --- | --- |
| Aldi 2018 ^1^ | Review (references checked) |
| Arancini 2021 ^2^ | UK data not separate for predictor analysis |
| Attar-Zadeh 2013 ^3^ | Not a study |
| Aubin 2009 ^4^ | Review (references checked) |
| Balmford 2011 ^5^ | UK data not separate for predictor analysis |
| Balmford 2014 ^6^ | UK data not separate for predictor analysis |
| Beard 2015 ^7^ | Outcomes - not cessation |
| Beard 2016 ^8^ | Outcomes - not cessation |
| Beard 2020 ^9^ | Outcomes - no OR or Beta correlation |
| Beard 2020 ^10^ | Outcomes - no OR or Beta correlation |
| Beard 2020 ^11^ | Outcomes - no OR or Beta correlation |
| Beard 2020 ^12^ | Outcomes - no OR or Beta correlation |
| Berg 2010 ^13^ | Outcomes - not cessation |
| Borland 2009 ^14^ | UK data not separate for predictor analysis |
| Borland 2010 ^15^ | UK data not separate for predictor analysis |
| Borland 2010 ^16^ | UK data not separate for predictor analysis |
| Borland 2012 ^17^ | UK data not separate for predictor analysis |
| Brose 2018 ^18^ | Review (references checked) |
| Brown 2014 ^19^ | Review of interventions for smoking cessation |
| Bryant 2011 ^20^ | Review (references checked) |
| Caleyachetty 2012 ^21^ | UK data not separate for predictor analysis |
| Ce 2015 ^22^ | Outcomes - not cessation |
| Cheung 2021 ^23^ | UK data not separate for predictor analysis |
| Clyde 2015 ^24^ | Outcomes - not cessation |
| Clyde 2018 ^25^ | Predictor not relevant |
| Cook 2014 ^26^ | Country - US |
| Cooper 2010 ^27^ | UK data not separate for predictor analysis |
| Cooper 2016 ^28^ | Population not MH (general) and UK data not separate for predictor analysis |
| Cooper 2016 ^29^ | Outcomes - not cessation (depressive symptoms), population not MGH (general), and UK data not separate for predictor analysis |
| Costello 2012 ^30^ | UK data not separate for predictor analysis |
| Doran 2019 ^31^ | UK data not separate for predictor analysis |
| Fidler 2011 ^32^ | Outcomes - not cessation |
| Fidler 2013 ^33^ | Outcomes - no OR or Beta correlation |
| Fix 2017 ^34^ | UK data not separate for predictor analysis |
| George 2012 ^35^ | Review (references checked) |
| Goldberg 2008 ^36^ | Outcome - no univariate/multivariate model of predictors, only a cross-tab of quitting and SZ medication |
| Guimond 2017 ^37^ | Outcomes - no OR or Beta correlation |
| Hakulinen 2015 ^38^ | No relevant predictors |
| Hall 2012 ^39^ | Not smoking cessation |
| Hall 2014 ^40^ | Suggests between country differences related to smoking cessation |
| Harmer 2013 ^41^ | Focussed on smoking relapse in pregnant women who had already quit smoking |
| Hawkins 2010 ^42^ | Smoking relapse |
| Herd 2009 ^43^ | Smoking relapse |
| Herd 2009 ^44^ | Smoking relapse |
| Ives 2008 ^45^ | Outcome (quit attempts rather than quitting) and setting (unclear whether hospital or not - no mention of hospital stay) |
| Khaled 2009 ^46^ | No prediction of cessation |
| Kock 2020 ^47^ | Does not contain anything on the predictors of cessation |
| Lacasse 2008 ^48^ | Outcomes - no OR or Beta correlation |
| Langdon 2013 ^49^ | Outcomes - not cessation |
| Lightfoot 2020 ^50^ | Review (references checked) |
| Lindson-Hawley 2016 ^51^ | Predictors not relevant |
| Martin 2020 ^52^ | Not examining predictors of cessation (examining predictors of smoking status) |
| NCT 2009 ^53^ | Protocol - no paper seems to be available |
| Nichols 2014 ^54^ | Protocol – no data. Looked up study and it does not involve prediction of quitting |
| Partos 2014 ^55^ | Data from 4 countries, UK data for predictors not presented separately |
| Poisson 2012 ^56^ | Data from 2 countries (NI and France), country as a predictor |
| Rae 2015 ^57^ | Outcomes - no OR or Beta correlation |
| Reid 2010 ^58^ | Data from 4 countries, UK data for predictors not presented separately |
| Siahpush 2009 ^59^ | Data from 4 countries, UK data for predictors not presented separately |
| Siahpush 2010 ^60^ | Data from 4 countries, UK data for predictors not presented separately |
| Smit 2011 ^61^ | Outcome (quit attempts rather than quitting) |
| Trainor 2017 ^62^ | Review (references checked) |
| Vaz 2017 ^63^ | Outcome - linear regression coefficient |
| Yong 2014 ^64^ | Data from 4 countries, UK data for predictors not presented separately |
| Yong 2018 ^65^ | Data from 4 countries, UK data for predictors not presented separately |
| Zhou 2014 ^66^ | Data from 5 countries, UK data for predictors not presented separately |

**Supplementary Table S2. Characteristics of included studies.**

| **Author, date** | **Study design** | **Country** | **Participants** | **Data set** | **Sample size** | **Time horizon of study** |
| --- | --- | --- | --- | --- | --- | --- |
| **General population UK** | | | | | | |
| Beard 2013^67^ | Longitudinal cohort | England | General population | Smoking Toolkit Study | 3149 | 6 months |
| Beenstock 2014^68^ | Randomised Controlled Trial | England | General population | ISRCTN22526020 | 422 | 6 months |
| Brose 2011^69^ | Intervention (service review) | England | Smoking cessation services | QuitManager | 126890 | 4 weeks |
| Brose 2012^70^ | Intervention (service review) | England | Smoking cessation services | QuitManager | 46237 and 12156 (separate analyses) | 4 weeks |
| Brose 2013^71^ | Intervention (service review) | England | Smoking cessation services | QuitManager | 167,487 | 4 weeks |
| Brown 2014^72^ | Retrospective cohort | England | General population | Smoking Toolkit Study | 5863 | 12 months |
| Fidler 2011^73^ | Longitudinal cohort | England | General population | Smoking Toolkit Study | 2257 | 6 months |
| Garnett 2020^74^ | Longitudinal cohort | England | General population | Smoking Toolkit Study | 2018 | 6 months |
| Gibson 2010^75^ | Longitudinal cohort | UK | General population | International Tobacco Control Four Country Survey | 2444 | 28 days |
| Graham 2010^76^ | Longitudinal cohort | UK | Pregnant women | Millennium Cohort Study | 4427 | 3 years |
| Hiscock 2011^77^ | Pooled data from two interventions (group and one to one support plus pharmacotherapy) | England and Scotland | Those aged 25-59 | NHS Stop Smoking Services (Glasgow, Nottingham, North Cumbria) | 2397 | 52 weeks |
| Hitchman 2015^78^ | Longitudinal cohort | UK | General population | An online longitudinal survey | 1643 | 1 year |
| Jackson 2019^79^ | Retrospective cohort | England | General population | Smoking Toolkit Study | 18929 | 12 months (previous) |
| Jackson 2019^80^ | Retrospective cohort | England | General population | Smoking Toolkit Study | 57522 | 12 months (previous) |
| Kale 2015^81^ | Randomised Controlled Trial | UK | General population | ESCAPE RCT | 4397 | 6 months |
| Kassim 2016^82^ | Intervention (service review) | England | English residents of Bangladeshi origin | NHS Stop Smoking Services (Tower Hamlets, London) | 324 | 4 weeks |
| Kotz 2009^83^ | Retrospective cohort | England | General population | Smoking Toolkit Study | 6950 | 12 months |
| Kotz 2014^84^ | Retrospective cohort | England | General population | Smoking Toolkit Study | 1560 | 6 months |
| McEwen 2009^85^ | Intervention (service review) | England | Smoking cessation services | NHS Stop Smoking Services (Merton, Sutton & Wandsworth; Redbridge & Waltham Forest) | 2626 | 4 weeks |
| Taggar 2015^86^ | Randomised Controlled Trial | England | General population | PORTSSS RCT | 2535 | 6 months |
| Walker 2018^87^ | Intervention (service review) | England | General population | Quit 51 smoking cessation services in Leicester, Lincoln, Sandwell, Solihull, Stoke, Surrey, East Sussex, Telford & Wreckin, Walsall, West Cheshire and Worcester) | 15640 (4 weeks) and 14273 (21 weeks) | 12 weeks |
| **Mental health UK and Canada** | | | | | | |
| Masuhara 2014^88^ | Intervention (service review) | Canada | Community Mental Health | Butt-Out Programme (Vancouver, Canada) service review | 90 | 26 weeks |
| Okoli 2011^89^ | Intervention (service review) | Canada | People with a mental illness and/or substance use disorder | Tobacco Dependence Clinic (Vancouver, Canada) | 258 | 26 weeks |
| Okoli 2014^90^ | Intervention (service review) | Canada | Community Mental Health / Substance Use - only those with psychiatric disorder extracted (i.e. not the univariate and multivariate models of predictors in the combined population) | Tobacco Dependence Clinic (Vancouver, Canada) | 522 | 28 weeks |
| Selby 2010^91^ | Intervention (service review, with matched pairs design [psychosis vs. no psychosis]) | Canada | People with a mental illness and/or those who have self-referred for nicotine dependence treatment | Centre for Addiction and Mental Health (CAMH) | 165 | End of treatment (not fixed duration) |
| Zawertailo 2015^92^ | Intervention (service review) | Canada | Smoking cessation | Smoking Treatment for Ontario Patients (STOP) | 5763 | 6 months |
| Zawertailo 2015^93^ | Intervention (service review) | Canada | Smoking cessation | NR | 1196 (3mo) and 846 (6mo) | 6 months |
| **Secondary care UK and Canada** | | | | | | |
| Khara 2015^94^ | Intervention (service review) | Canada | Secondary care | Vancouver General Hospital Smoking Cessation Clinic | 117 | 8 months and 1 year |
| Salman 2020^95^ | Longitudinal cohort | UK | Cardiac rehab | National Audit of Cardiac Rehabilitation database | 3290 | 9 weeks (pre-post cardiac rehab, mean duration) |

CAMH, Centre for Addiction and Mental Health; NHS, National Health Service; NR, not reported; RCT, randomised controlled trial; STOP, Smoking Treatment for Ontario Patients; UK, United Kingdom.

### Supplementary Table S3: Participant baseline characteristics of included studies

| **Author, date** | **Age (Mean (SD))** | **Sex** | **Socio-economic status** | **Reason for admission (if relevant)** | **Cigarettes per day (mean (SD))** | **Number of previous quit attempts** | **Nicotine dependence** |
| --- | --- | --- | --- | --- | --- | --- | --- |
| **General population UK** | | | | | | |  |
| Beard 2013^67^ | 40.4 (16.09) | 51% male | 15.3% AB, 25.2% C1, 24.6% C2, 21.9% D,  13.0% E | NR | 13.3 (8.43) | NR | 68% reported having a cigarette <30 mins of waking |
| Beenstock 2014^68^ | Median 49 (IQR 40 to 57) | 50.2% male | 49.5% had post-school qualifications (47.8% did not) | NR | NR | NR | Median FTND score: 6 (IQR 4 to 7) |
| Brose 2011^69^ | NR | NR | NR | NR | NR | NR | NR |
| Brose 2012^70^ | NR | NR | Mean IMD score 23.2 (SD 8.6, range 11.4 to 45.3) | NR | NR | NR | Mean HSI 3.4 (SD 1.5) of subsample with HSI data |
| Brose 2013^71^ | NR | 52.4% female | 13.8% managerial / professional, 7.2% intermediate, 22.4% routine / manual, 11.9% retired, 4.5% student, 13.0% long-term unemployed, 5.6% home carer, 1.3% in prison | NR | NR | NR | NR |
| Brown 2014^72^ | NR | NR | NR | NR | NR | NR | NR |
| Fidler 2011^73^ | 46.3 (15.2) | 41.5% male | 31.3% A, B, C1 68.7% C2, D, E | NR | NR | NR | NR |
| Garnett 2020^74^ | 46.0 (15.96) | 54.5% female | 12.9% AB, 23.0% C1, 21.7% C2, 17.3% D, 25.1% E | NR | 13.1 (8.89) | In past 6 months:  1: 69.5%, 2: 21.3%, ≥3: 9.2% | NR |
| Gibson 2010^75^ | Median 44 (IQR 34 to 55) | 44% male | NR | NR | Median 18 (IQR 12 to 20) | Median 3 (IQR 2 to 5) | Mean HSI 2.5 (SD 1.5) |
| Graham 2010^76^ | NR | 100% female | Current NS-SEC:  Highest 31.3%, intermediate 23.4%, lowest 41.3%, economically inactive 3.9% | NR | NR | NR | NR |
| Hiscock 2011^77^ | NR | NR | NR | NR | NR | NR | NR |
| Hitchman 2015^78^ | 9.98% aged 18-24, 27.94% aged 25-39, 32.14% aged 40-54, 29.95%% aged ≥55 | 44.3% male | Income:  10.96% high, 39.01% medium, 34.75% low | NR | NR | NR | NR |
| Jackson 2019^80^ | 14.3% aged 16-24, 16.7% aged 25-34, 16.6% aged 35-44, 17.4% aged 45-54, 21.0% aged ≥65 | 51.0% female | 27.1% AB, 27.5% C1, 21.8% C2, 15.0% D, 8.6% E | NR | NR | NR | NR |
| Jackson 2019^79^ | 20.4% aged 16-24, 23.3% aged 25-34,  20.2% aged 35-44, 15.8% aged 45-54, 11.7% aged 55-64, 8.7% aged ≥65 | 52.0% female | 62.9% grades C2, D, E | NR | NR | In past year:  1 – 65.3% 2 – 21.2%  3 – 7.4%  ≥4 – 6.1% | NR |
| Kale 2015^81^ | NR | NR | NR | NR | NR | NR | NR |
| Kassim 2016^82^ | 45.59 (13.83) | 10% female | Mean IMD small area score 56.98 (SD 5.37);  Mean IMD postcode score 50.98 (SD 11.35) | NR | 12.71 (7.08) | Mean 1.34 (SD 1.59) | NR |
| Kotz 2009^83^ | 40.6 (16.4) | 52.2% male | 16.8% AB, 25.6% C1, 23.8% C2, 22.2% D, 11.6% E | NR | 13.4 (8.5) |  | Mean FTND score 2.9 (SD 2.4) |
| Kotz 2014^84^ | 46.5 (15.7) | 55.8% female | 11.0% AB, 20.9% C1, 22.0% C2, 17.5% D, 28.7% E | NR | NR | 0 – 69.2%, 1 – 22.5%, 2 – 8.3% | NR |
| McEwen 2009^85^ | 45 (13.8) | 59% female | Education >16 years of age: 41%;  In paid employment 46% | NR | 21.4 (10.1) | 1 – 27%, >1 – 54% | Mean FTND 5.7 (SD 2.2) |
| Taggar 2015^86^ | Median 38 (IQR 28 to 50) | 53.5% female | Median IMD score 23.1 (IQR 13.5 to 37.1) | NR | NR | NR | Mean HSI score 3 (SD 1.6) |
| Walker 2018^87^ | 44.8 (15.6) | 47.6% male | Mean deprivation index 6.54 (SD 2.73) | NR | NR | NR | Mean FTND 4.68 (SD 2.24) |
| **Mental health UK and Canada** | | | | | | |  |
| Masuhara 2014^88^ | 47.1 (11.5 years) | 53.7% male | NR | NR | NR | ≤5 – 79.9% >5 – 20.1% | Mean FTND 6.0 (SD 2.1) |
| Okoli 2011^89^ | 48.6 (11.0) | 62.8% male | NR | NR | NR | NR | NR |
| Okoli 2014^90^ | 48.0 (11.1) | 59.2% male | NR | NR | 20.4 (10.3) | NR | Mean FTND 6.0 (SD 2.1) |
| Selby 2010^91^ | 46.2 (9.8) | 61.8% male | NR | NR | 25.9 (12.9) | 0 – 9.1%  1-4 – 57.0%  ≥5 – 33.9% | Mean FTND 6.7 (SD 1.9) |
| Zawertailo 2015^92^ | 43.5 (12.5) | 58.5% female | Household income:  ≤$20,000 29.5%, $20,000-40,000 28.1%,  $40,000-80,000 30.8%,  >$80,000 11.7% | NR | Heavy smoking ≥20 cigarettes per day, 68.2% | 0 – 6.8% 1-5 – 68.7% ≥6 – 24.5% | HSI score:  Low 10.4%, Moderate 47.6%, High 42.0% |
| Zawertailo 2015^93^ | 46.24 (13.12) | 62.8% male | Annual household income:  ≤$40,000 80.3%, >$40,000 18.7%  Education:  Less than secondary 25.6%, High school diploma 18.9%, Post-secondary 55.5%  Employment status:  Employed 26.4%, Unemployed 26.6%, Student/retired 9.8%, Disability 37.2% | NR | 20.96 (11.95) | NR | Time to first cigarette:  <5 min 47.0%, 6-30 min 32.9%, 31-60 min 7.6%, >60 min 6.5% |
| **Secondary care UK and Canada** | | | | | | |  |
| Khara 2015^94^ | 58.5 (10.5) | 66.4% male | Income source:  Disability / social assistance 30.2%, State pension 31.0, Earned income 38.8% | Referral source:  Cardiology 79.5%,  Respirology 10.3%, Other sources 10.3% | 15.9 (8.0) | NR | Mean FTND 4.2 (2.4) |
| Salman 2020^95^ | Smokers: 58.59 (10.49), Quitters: 57.63 (10.36) | NR | Mean IMD decile:  Smokers 4.86 (SD 2.90)  Quitters 4.89 (SD 2.90 | NR | NR | NR | NR |

### Supplementary Table S4: Outcome measurement and analyses in included studies

| **Author, date** | **Measure of cessation** | **Cessation timepoint** | **Quit success** | **Model type** | **ORs captured (yes/no)** | **Beta Coefficients (yes/no)** |
| --- | --- | --- | --- | --- | --- | --- |
| **General population UK** | | | | | | |
| Beard 2013^67^ | 4-week point-prevalence abstinence, self-reported | Between 4 weeks and 6 months | 9% | Logistic regression | Yes | No |
| Beenstock 2014^68^ | Russell standard abstinence, allowing for 2-week grade period from quit date, verified by CO (<10ppm) | 4 weeks and 6 months | 44.2% at 4 weeks, 18.8% at 6 months | Logistic regression | Yes | No |
| Brose 2011^69^ | 2-week point-prevalent abstinence verified by CO (<=10ppm) | Between 2 and 4 weeks | Mean 36.0% (range 3.8% to 56.4%) | Multiple logistic regression | Yes | No |
| Brose 2012^70^ | 2-week point-prevalent abstinence verified by CO (<=10ppm) | Between 2 and 4 weeks | 39% CO validated abstinence | Multi-level model (3 levels) | Yes | No |
| Brose 2013^71^ | 2-week point-prevalent abstinence verified by CO (<=10ppm) | Between 2 and 4 weeks | 34.7% CO validated | Multi-level model (3 levels) | Yes | No |
| Brown 2014^72^ | Self-reported abstinence | Up to 12 months | 14% | Logistic regression | Yes | No |
| Fidler 2011^73^ | Self-reported continuous abstinence | Between 4 weeks and 6 months | 7.1% of sample, 18% of those who had made a quit attempt | Logistic regression | Yes | No |
| Garnett 2020^74^ | Self-reported smoking status (successful/unsuccessful) | 6 months | NR | GLM | Yes | No |
| Gibson 2010^75^ | Self-reported abstinence | 28 days | NR | Logistic regression | Yes | No |
| Graham 2010^76^ | Self-reported smoking status | In pregnancy | 36% quit during pregnancy, 15% were abstinent 9 months post-partum (43% of the 36%) | Logistic regression | Yes | No |
| Hiscock 2011^77^ | CO validated continuous abstinence | 52 weeks | 9.6% | Logistic regression | Yes | No |
| Hitchman 2015^78^ | Self-reported smoking status | 1 year | NR | Logistic regression | Yes | No |
| Jackson 2019^80^ | Self-reported abstinence | Start of most recent quit attempt to time of survey (within last 12 months) | NR | Logistic regression | Yes | No |
| Jackson 2019^79^ | Self-reported continuous abstinence | Start of most recent quit attempt to time of survey (within last 12 months) | 15.7% | Multiple logistic regression | Yes | No |
| Kale 2015^81^ | Self-reported continuous abstinence (prior 3 months) | 6 months | 4.2% | Logistic regression | Yes | No |
| Kassim 2016^82^ | Validated smoking cessation (method of validation not specified, although probably CO given context is service) | 4 weeks | 68.8% | Logistic regression | Yes | No |
| Kotz 2009^83^ | Self-reported continuous abstinence | 12 months | 15.7% | Multiple logistic regression | Yes | No |
| Kotz 2014^84^ | Self-reported continuous abstinence | 6 months | 23.0% | Multiple logistic regression | Yes | No |
| McEwen 2009^85^ | 2-week point-prevalent abstinence verified by CO (<=10ppm) | Between 2 and 4 weeks | 41% | Forward stepwise multiple logistic regression | Yes | No |
| Taggar 2015^86^ | Self-reported abstinence | 6 months (from quit date) | 18.5% | Logistic regression | Yes | No |
| Walker 2018^87^ | CO-validated abstinence at 4 weeks, self-reported abstinence at 12 weeks | 4 and 12 weeks | Mean quit rate:  56.0% at 4 weeks,  28.8% at 12 weeks | Generalised linear mixed model (stepwise) | Yes | Yes |
| **Mental health UK and Canada** | | | | | | |
| Masuhara 2014^88^ | 7-day point-prevalent abstinence verified by CO (<=8ppm) | Any time from 12 to 26 weeks | 26.7% among programme completers | Logistic regression (stepwise) | Yes | No |
| Okoli 2011^89^ | 7-day point-prevalent abstinence verified by CO (<=8ppm) | Any time from 8 to 26 weeks | 32.2% | 2-step logistic model | Yes | No |
| Okoli 2014^90^ | 7-day point-prevalent abstinence verified by CO (<=8ppm) | Any time from 8 to 28 weeks | 40.6% of programme completers  (33.1% of all participants) | Logistic regression (2-step?) | Yes | No |
| Selby 2010^91^ | 7-day point-prevalent abstinence | End of treatment (not fixed duration) | 20.6% | Logistic regression | Yes | No |
| Zawertailo 2015^92^ | Self-reported quitting, self-reported abstinence and 30-day point-prevalent abstinence | 6 months | NR | Logistic regression | Yes | No |
| Zawertailo 2015^93^ | Self-reported 7-day point-prevalence abstinence | 3 months and 6 months | NR | Logistic regression | Yes | No |
| **Secondary care UK and Canada** | | | | | | |
| Khara 2015^94^ | 7-day point-prevalent abstinence verified by CO (<=8ppm) | Last recorded data collection point (last follow-up visit) | 1 month 24.8%,  6 months 12.8% | 2-step logistic regression | Yes | No |
| Salman 2020^95^ | Unclear. Possibly self-report. | 9 weeks (pre-post cardiac rehab, mean duration 9 weeks) | 37.6% | Logistic regression | Yes | Yes |

**Supplementary Table S5. Relationships between covariates and quit success.**

| **Covariate** | **Measure or category** | **Significantly lower odds of quitting** | **No evidence of a relationship** | **Significantly greater odds of quitting** |
| --- | --- | --- | --- | --- |
| **Demographics** | | | | |
| Age | Older |  | ^68,73,74,76,81,82^ ^88,90^ ^94,95^ | ^70,71,77,85,87^ |
| Sex | Male | ^68,85^ | ^71,73,78,81,82,87^ ^88,90^ | ^70,74,77^ ^91^ ^94^ |
| Ethnicity | Black |  | ^81^ | ^77^ |
| Socio-economic conditions | Socio-economic status (high) |  |  | ^70,73,74,76,77,81,83^ |
|  | IMD (low deprivation) |  | ^70^ | ^87^ |
|  | Household income (high) |  |  | ^76,78^ |
|  | Education (highest qualification) |  | ^78,81,85^ | ^68^ |
|  | Age of leaving education (older) |  |  | ^76^ |
|  | Client pays for prescription |  | ^85^ | ^70,71^ |
|  | Reading level (high) |  |  | ^81^ |
|  | Childhood NS-SEC socio-economic classification of mother (higher) |  |  | ^76^ |
|  | Employment status |  | ^85^ ^94,95^ |  |
|  | Social housing (vs. other housing) | ^80^ |  |  |
| Relationship, cohabitation and social network | Married, living with partner | ^85^ | ^76,81^ | ^95^ |
|  | Other household smokers |  | ^77,85^ |  |
|  | Social support from family/friends (more support) |  | ^77,81^ | ^88^ |
| **Health and healthcare setting** | | | | |
| Physical health | Cardiovascular risk (low) |  |  | ^95^ |
|  | Comorbidities (more) | ^95^ | ^94^ |  |
|  | Referral source (cardiology or respirology vs. other) |  | ^94^ |  |
| Mental health | Primary mental health diagnosis (vs. no diagnosis) | ^90^ |  |  |
|  | Primary mental health diagnosis (vs. each other) |  | ^88-91^ |  |
|  | Anxiety score (severe) |  | ^95^ |  |
|  | Depression score (severe) |  | ^88,93^ ^95^ |  |
|  | History of depression (vs. no history) | ^92^ |  |  |
|  | History of psychiatric disorder (vs. no history) |  | ^94^ |  |
|  | History of substance abuse | ^89,90^ | ^88^ |  |
|  | Co-occurring psychiatric and substance use disorder |  | ^94^ |  |
| **Tobacco smoking variables** | | | | |
| Current and previous tobacco consumption, and nicotine dependence | Daily cigarette consumption (more/increasing) | ^76^ | ^74^ ^88,90^ ^94^ |  |
|  | CO level at baseline (higher / increasing) | ^89^ | ^90^ ^94^ |  |
|  | Age at smoking initiation (older) |  | ^81^ ^88,90^ ^94^ |  |
|  | Nicotine dependence (higher / increasing) | ^68,70,73,74,78,81,85-87^ ^88,90^ | ^77^ ^89,91^ ^94^ |  |
|  | Most difficult situation not to smoke (feeling urge to smoke) | ^81^ |  |  |
|  | Most difficult situation not to smoke (socialising, first thing in the morning, angry / frustrated, relaxing, other) |  | ^81^ |  |
|  | Enjoyment of smoking (high / increasing) |  | ^73^ |  |
| Motivation | Determination to quit (vs. not) | ^77^ |  |  |
|  | Motivation to quit (vs. not motivated) | ^78^ |  |  |
|  | Importance of quitting (high / increasing) |  | ^88,90^ ^94^ |  |
|  | Confidence in quitting |  | ^88,90^ ^94^ |  |
|  | Readiness to quit (higher) |  | ^81^ ^88^ ^94^ |  |
|  | Reason for quitting |  | ^81^ |  |
|  | Main advantage of quitting |  | ^81^ |  |
|  | Main disadvantage of quitting |  | ^81^ |  |
| Quitting characteristics | Past serious quit attempt (vs. none) |  | ^69,85^ |  |
|  | Number of previous quit attempts (higher) |  | ^88^ | ^74,81^ |
|  | Duration of previous quit attempts (longer) |  | ^94^ | ^68^ ^90^ |
|  | Time since start of most recent quit attempt (longer) |  | ^74^ |  |
| **Intervention characteristics** | | | | |
| Reduction in amount smoked and/or temporary abstinence prior to quitting | Reduction in smoking prior to quitting |  | ^68^ | ^67,74^ |
|  | Reduction in smoking with NRT prior to quitting |  |  | ^67^ |
|  | Temporary abstinence with NRT prior to quitting |  |  | ^67^ |
| Behavioural support type, setting and mode of contact | Smoking cessation support from a clinic or helpline (vs. no support) |  |  | ^75^ |
|  | Face-to-face support |  | ^79^ |  |
|  | Telephone support | ^79^ |  |  |
|  | Written self-help materials |  | ^79^ |  |
|  | Support from websites |  |  | ^79^ |
|  | Support from a doctor or other health professional (vs. no support) |  | ^75^ |  |
|  | Support from pharmacotherapy plus clinic or health professional (vs. no support) |  |  | ^75^ |
|  | Support in primary care setting (vs. specialist clinic) | ^69,70^ |  |  |
|  | Support from specialist clinics |  | ^71^ |  |
|  | Support in pharmacy setting |  | ^69^ |  |
|  | Community support |  |  | ^85,87^ |
|  | Group support |  | ^91^ | ^69,71^ |
|  | One-to-one support |  | ^72^ |  |
|  | Drop-in support | ^69^ |  |  |
| Tobacco dependence treatment duration and number of contacts | Greater number treatment sessions / contacts |  | ^88^ ^94^ | ^77^  ^89-91^ |
|  | Length of time in programme |  | ^94^ |  |
|  | Practitioner experience (greater number of treatment episodes delivered) |  | ^70^ |  |
| Pharmacological aids | Pharmacological aids in general (vs. no aids) |  |  | ^75,84^ |
|  | Combination therapy (vs. a single pharmacological aid) |  | ^88,90,91^ |  |
|  | NRT | ^79,84^ | ^90^ | ^69,70,79^ |
|  | Combination NRT |  |  | ^70,82^ |
|  | Varenicline |  | ^90^ | ^69,70,79,87^ |
|  | Bupropion |  | ^69,79^ ^90^ | ^70,85^ |
|  | E-cigarettes | ^78^ |  | ^72,78,79^ |
|  | Compliance with pharmacotherapy |  |  | ^77^ |
| Interaction terms | Treatment x setting |  | ^87^ |  |
|  | Treatment x FTND |  | ^87^ |  |

CO, carbon monoxide; FTND, Fagerstrom Test for Nicotine Dependence; IMD, Indices of Multiple Deprivation; NRT, nicotine replacement therapy; NS-SEC, The National Statistics Socio-economic classification

Key: Black = UK general population; blue = Mental health setting UK and Canada; orange = Secondary care setting UK and Canada

### Supplementary Table S6: Covariates of quitting outcomes – full results summary

| **Covariate** | **Setting** | **Significantly lower odds of cessation** | **No evidence of a relationship** | **Significantly greater odds of cessation** |
| --- | --- | --- | --- | --- |
| **Demographics** | | | | |
| Age (older) | General population UK |  | - Odds of cessation at 6 months were not greater for 25-34, 35-44, 45-54 or ≥65 years age groups than for 16-24 year age group, in adjusted and unadjusted analyses ^74^ - Odds of cessation were not greater for 35-42, 43-50 or 51-59 year olds than for 25-34 year olds in adjusted analyses of patients from England ^77^ - Odds of cessation at 1 year were not greater for 38-47 year olds than for 25-37 year olds in adjusted analyses of patients from one-to-one clinics in Glasgow ^77^ - There was no impact of age (as a continuous variable) on cessation at 6 months in either univariate or multivariate analyses ^81^ - There was no impact of age (in 1-year increments) on cessation at 6 months in an adjusted analysis ^82^ - There was no impact of age (in 1-year increments) on cessation at 4 weeks to 6 months in univariate analysis ^73^ - There was no impact of age (as a continuous variable) on cessation at either 4 weeks or 6 months in adjusted analyses ^68^ | - Odds of cessation at 1 year were significantly greater among 48-59 year olds than among 25-37 year olds (OR 5.8, 95% 1.1 to 30.2) in adjusted analyses of patients from one-to-one clinics in Glasgow ^77^ - Odds of cessation at 4 weeks were significantly greater among 20-39 year olds (OR 1.77, 95% CI 1.51 to 2.08), 40-59 year olds (OR 2.05, 95% CI 1.74 to 2.40) and those aged ≥60 years (OR 2.41, 95% CI 2.03 to 2.86) than among 13-19 year olds in adjusted analyses ^87^ - There was a significant impact of age (in 1-year increments) on cessation at 4 weeks in an adjusted analysis (OR 1.02, 95% 1.02 to 1.02), and in a sub-population of only those with HSI data (OR 1.02, 95% CI 1.02 to 1.03) ^70^ - There was a significant impact of age (in 10-year increments) on cessation at 4 weeks in an adjusted analysis (OR 1.2, 95% 1.19 to 1.21) ^71^ - There was a significant impact of age (continuous) on cessation at 4 weeks in unadjusted and adjusted analyses (all OR 1.02, p<0.001) ^71^ |
|  | Mental health UK Canada |  | - There was no impact of age (as a continuous variable) on cessation at 12-26 weeks in unadjusted or adjusted analyses ^88^ - There was no impact of age (as a continuous variable) on cessation at 8-28 weeks in unadjusted analysis ^90^ |  |
|  | Secondary care UK Canada |  | - There was no impact of age (continuous) on cessation at most recent follow-up in an adjusted analysis (OR 1.0, 95% 1.0 to 1.0) ^94^ - There was no impact of age (as a continuous variable) on cessation at around 9 weeks in unadjusted analysis ^94^ |  |
| Smoking cessation in pregnant women – moderating effect of age at first live birth | General population UK |  | - The odds of cessation over the duration of pregnancy were not significantly different for the 25-29, 20-24 and 14-19 years age groups compared with the ≥30 years age group in a mutually adjusted analysis and a mutually adjusted analysis including cigarette consumption ^76^ | - The odds of cessation over the duration of pregnancy were significantly lower in the 20-24 (OR 0.46, 95% CI 0.38 to 0.57) and 14-19 (OR 0.42, 95% CI 0.34 to 0.52) years age groups compared with the ≥30 age group, but not significantly different between the 25-29 and ≥30 age groups (OR 0.85, 95% CI 0.67 to 1.08) in an unadjusted analysis ^76^ |
|  | Mental health UK Canada | No data | No data | No data |
|  | Secondary care UK Canada | No data | No data | No data |
| Sex (male) | General population UK | - The odds of cessation at 4 weeks were significantly higher in females than males in a univariate analysis (OR 1.18. p=0.044), adjusted analysis including all variables reaching significance in the univariate analyses (OR 1.3, p=0.009) and adjusted analysis including all variables reaching significance in the univariate analyses alongside choice of NRT product (OR 1.39, p=0.003) ^85^ - The odds of cessation at 4 weeks were significantly lower in males than females in analyses adjusted for all other variables including the Consideration of Future Consequences (CFCS) scale (OR 0.72, 95% CI 0.52 to 0.99), CFCS future subscale (OR 0.71, 95% CI 0.52 to 0.98) and CFCS immediate subscale (OR 0.72, 95% CI 0.52 to 0.99) ^68^ | - The odds of cessation at 1 year were not significantly different in males (OR 0.9, 95% CI 0.3 to 2.8) than females in adjusted analyses of one-to-one services in Glasgow ^77^ - The odds of cessation at 1 year were not significantly different in females (OR 1.22, 95% CI 0.89 to 1.67) than males in adjusted analyses ^78^ - The odds of cessation at 6 months were not significantly different in females than males in univariate (OR 1.02, 95% CI 0.74 to 1.40) and multivariate (OR 0.81, 95% CI 0.56 to 1.18) analyses ^81^ - The odds of cessation at 4 weeks were not significantly different in females (OR 1.28, 95% CI 0.54 to 3.06) than males in adjusted analyses ^82^ - The odds of cessation at 4 weeks were not significantly different in females (OR 0.96, 95% CI 0.90 to 1.02) than males in adjusted analyses ^87^ - The odds of cessation at 4 weeks to 6 months were not significantly different in females (OR 0.66, 95% CI 0.43 to 1.01) than males in an adjusted analysis ^73^ - The odds of cessation at 4 weeks were not significantly different in males (OR 1.00, 95% CI 0.98, 1.02) than females in an adjusted analysis ^71^ - The odds of cessation at 6 months were not significantly different in males than females in analyses adjusted for all other variables including the Consideration of Future Consequences (CFCS) scale (OR 0.71, 95% CI 0.47 to 1.07), CFCS future subscale (OR 0.70, 95% CI 0.46 to 1.06) and CFCS immediate subscale (OR 0.71, 95% CI 0.47 to 1.07) ^68^ | - The odds of cessation at 1 year were significantly higher in males (OR 1.29, 95% CI 1.05 to 1.58) than females in unadjusted and adjusted analyses ^74^ - The odds of cessation at 6 months were significantly higher in males (OR 1.5, 95% CI 1.1 to 2.1) than females in adjusted analyses of smoking cessation services in England ^77^ - The odds of cessation at 4 weeks to 6 months were significantly lower in females (OR 0.62, 95% CI 0.41 to 0.94) than males in a univariate analysis ^73^ - The odds of cessation at 4 weeks were significantly higher in males (OR 1.11, 95% CI 1.07 to 1.16) than females in an adjusted analysis, as well as in a sub-sample of only those with HSI data (OR 1.16, 95% CI 1.07 to 1.26) ^70^ |
|  | Mental health UK Canada |  | - The odds of cessation at 12 to 26 weeks were not significantly different in females (OR 1.4, 95% CI 0.6 to 3.7) than males in univariate analysis ^88^ - The odds of cessation at 8 to 28 weeks were not significantly different in females (OR 0.79, 95% CI 0.56 to 1.13) than males in an unadjusted analysis, as well as in an adjusted analysis of a substance use disorder sub-population (OR 0.53, 95% CI 0.21 to 1.32) ^90^ | - The odds of cessation at end of treatment were significantly higher in males (OR 2.75, 95% CI 1.01 to 7.49) than females in an adjusted analysis ^91^ |
|  | Secondary care UK Canada |  |  | - The odds of cessation at most recent follow-up were lower among females than males in unadjusted (OR 0.3, 95% CI 0.1 to 0.8) and adjusted (OR 0.3, 95% CI 0.1 to 1.0) analyses ^94^ |
| Ethnicity | General population UK |  | - The odds of cessation at 1 year were not significantly different in those who mentioned Asian ethnicity (OR 0.6, 95% CI 0.1 to 4.9) or other/unknown ethnicity (OR 0.2, 95% CI 0.00 to 1.5) compared with White British in an adjusted analysis ^77^ - The odds of cessation at 6 months were not significantly different among those with Non-White compared with White ethnicity in univariate (OR 0.53, 95% CI 0.17 to 1.71) and multivariate (OR 0.4, 95% CI 0.08 to 2.06) analyses ^81^ | - The odds of cessation were significantly higher among those who mentioned Black ethnicity (OR 6.9, 95% CI 2.1 to 23.2) compared with White British ethnicity in an adjusted analysis ^77^ |
|  | Mental health UK Canada | No data | No data | No data |
|  | Secondary care UK Canada | No data | No data | No data |
| Socio-economic status (high) | General population UK |  | - The odds of cessation at 6 months were not significantly different for those in grades C1, C2 and D in both unadjusted and adjusted analyses ^74^ - The odds of cessation at 1 year were not significantly different in those with 2 (OR 1.6, 95% CI 0.8 to 3.0) and 3 (OR 1.1, 0.6 to 2.2) affluence indicators compared with 0-1, in England smoking cessation services, nor between those who had 2-4 (OR 2.0, 95% CI 0.4 to 9.8) and all affluence indicators (OR 5.1, 95% CI 0.8 to 31.2) compared with those who had 0 or 1 affluence indicator in Glasgow one-to-one services, in adjusted analyses ^77^ - There was no significant difference in odds of cessation at 6 months for those in social grades 1 and 5 (most deprived) compared with those in category 0 (least deprived) in both univariate (OR 0.82, 95% CI 0.57 to 1.19; OR 0.44, 95% CI 0.14 to 1.41, respectively) and multivariate (OR 0.89, 95% CI 0.59 to 1.33; OR 1.34, 95% CI 0.34 to 5.24, respectively) analyses ^77^ - There was no significant difference in odds of cessation at 1 year between those in social grade C1 (OR 0.79, 95% CI 0.59 to 1.05) and those in AB in an adjusted analysis ^83^ - There was no difference in the odds of cessation at 4 weeks between those who were students (OR 0.94, 95% CI 0.83 to 1.05), retired (OR 1.08, 95% CI 0.99 to 1.18), home carers (OR 0.98, 95% CI 0.90 to 1.08) and those with uncodable data (OR 1.03, 95% CI 0.93 to 1.13) compared with those in routine and manual occupations in adjusted analyses, or between any other occupational category and those in routine and manual occupations in an adjusted analysis of a sub-sample including only those with HSI data ^70^ | - Those in social grade E had significantly lower odds of cessation at 6 months than those in AB, in both unadjusted (OR 0.48, 95% CI 0.34 to 0.69) and adjusted (OR 0.65, 95 CI 0.45 to 0.69) analyses ^74^ - Those who had 4 affluence indicators (OR 2.3, 95% CI 1.3 to 4.1) and those who had all affluence indicators (OR 2.3, 95% CI 1.2 to 4.4) had significantly higher odds of cessation at 1 year than those with 0-1 affluence indicators in English smoking cessation services in an adjusted analysis ^77^ - Those in social deprivation categories 2 and 3 had significantly lower odds of cessation at 6 months than those in category 0 (least deprived) in both univariate (OR 0.47, 95% CI 0.30 to 0.74; OR 0.26, 95% CI 0.12 to 0.57, respectively) and multivariate (OR 0.59, 95% CI 0.35 to 0.98; OR 0.3, 95% CI 0.11 to 0.79, respectively), whereas those in social deprivation category 4 had lower odds of cessation than those in category 0 in univariate (OR 0.43, 95% CI 0.20 to 0.93) but not multivariate (OR 0.77, 95% CI 0.30 to 1.97) analyses ^81^ - Those in grades C2 (OR 0.67, 95% CI 0.49 to 0.90), D (OR 0.68, 95% CI 0.50 to 0.93) and E (OR 0.5, 95% CI 0.33 to 0.74) had significantly lower odds of cessation at 1 year than those in grades AB in adjusted analysis ^83^ - Those in lower social grades had significantly lower odds of cessation at 4 weeks to 6 months per grade increase in univariate (OR 0.82, 95% CI 0.71 to 0.96) and adjusted (OR 0.85, 95% CI 0.73 to 1.00) analyses ^73^ - The odds of cessation at 4 weeks were significantly lower among those who were long-term unemployed (OR 0.81, 95% CI 0.75 to 0.87) and those who were sick/disabled and unable to work (OR 0.75, 95% CI 0.68 to 0.82), and significantly higher among those with managerial and professional occupations (OR 1.14, 95% CI 1.06 to 1.23) , those in intermediate occupations (OR 1.13, 95% CI 1.04 to 1.23) and those in prison (OR 2.01, 95% CI 1.68 to 2.39) compared with those in routine and manual occupations, in an adjusted analysis ^70^ - Pregnant women in intermediate occupations (OR 0.6, 95% CI 0.48 to 0.75), routine and manual occupations (OR 0.36, 95% CI 0.3 to 0.43) and those who were long-term unemployed (OR 0.19, 95% CI 0.13 to 0.28) compared with those in managerial and professional occupations in unadjusted analyses and mutually adjusted analyses including cigarette consumption (OR 0.76, 95% CI 0.58 to 0.98; OR 0.63, 95% CI 0.48 to 0.81; OR 0.35, 95% CI 0.23 to 0.54, respectively), but was only significantly lower than managerial and professional occupations for those in routine and manual occupations (OR 0.36, 95% CI 0.52 to 0.84) and those who were long-term unemployed (OR 0.38, 95% CI 0.25 to 0.56) in mutually adjusted analyses, with no significant difference for intermediate occupations (OR 0.81, 95% CI 0.64 to 1.02) ^76^ |
|  | Mental health UK Canada | No data | No data | No data |
|  | Secondary care UK Canada | No data | No data | No data |
| Indices of multiple deprivation (IMD) | General population UK |  |  | - The odds of cessation at 4 weeks significantly decreased as IMD (continuous) increased (OR 0.95, 95% CI 0.94 to 0.97) in an adjusted analysis ^87^ |
|  | Mental health UK Canada | No data | No data | No data |
|  | Secondary care UK Canada | No data | No data | No data |
| Service-level IMD score | General population UK |  | - The odds of cessation at 4 weeks were not significantly impacted by service-level IMD (per unit increase) in an adjusted analysis (OR 0.98, 95% CI 0.96 to 1.01), nor a sub-sample of only those who had HSI data (OR 0.99, 95% CI 0.96 to 1.03) ^70^ |  |
|  | Mental health UK Canada | No data | No data | No data |
|  | Secondary care UK Canada | No data | No data | No data |
| Household income | General population UK |  | - The odds of cessation at the end of pregnancy were not significantly different among women with a household income of £22000-33000 (OR 0.96, 95% CI 0.71 to 1.29) in a mutually adjusted analysis, and among those with a household income of £22000-33000 (OR 1.08, 95% CI 0.79 to 1.47) and £11000-22000 (OR 0.82, 95% CI 0.59 to 1.13) in a mutually adjusted analysis including cigarette consumption ^76^ - The odds of cessation were not significantly different among those with a medium income (OR 1.21, 95% CI 0.83 to 1.78) or who gave no answer (OR 1.4, 95% CI 0.82 to 2.39) than those with a low income in an adjusted analysis ^78^ | - The odds of cessation at the end of pregnancy were significantly lower among pregnant women with a household income of £22000-33000, £11000-22000 and £0-11000 than among those with a household income of ≥£33000 in an unadjusted analysis (OR 0.72, 95% CI 0.55 to 0.95; OR 0.45, 95% CI 0.35 to 0.58; OR 0.30, 95% CI 0.23 to 0.40, respectively), among those with a household income of £11000-22000 and £0-11000 in a mutually adjusted analysis (OR 0.72, 95% CI 0.53 to 0.96; OR 0.52, 95% CI 0.37 to 0.74, respectively) and among those with a household income of £0-11000 in a mutually adjusted analysis including cigarette consumption (OR 0.61, 95% CI 0.42 to 0.88) ^76^ - The odds of cessation at 1 year were significantly higher among those with a high income (OR 1.63, 95% CI 1.00 to 2.66) than a low income in an adjusted analysis ^78^ |
|  | Mental health UK Canada | No data | No data | No data |
|  | Secondary care UK Canada | No data | No data | No data |
| Education – highest qualification | General population UK |  | - There was no significant difference between the odds of cessation at 1 year among those with medium (OR 1.04, 95% CI 0.73 to 1.50) and high (OR 0.82, 95% CI 0.54 to 1.26) education compared with those with low education in an adjusted analysis ^78^ - There was no significant difference between the odds of cessation at 6 months among those who had A-level education or higher than those with education to below A-level in univariate (OR 1.25, 95% CI 0.91 to 1.71) or multivariate (OR 0.56, 95% CI 0.32 to 1.00) analyses ^81^ - There was no significant difference in the odds of cessation at 4 weeks between those who had no qualifications and those who had GCSE or above (OR 1, p=0.98) in an unadjusted analysis ^85^ - The odds of cessation at 6 months were not significantly different in those with than without post-school qualifications in analyses adjusted for all other variables including the Consideration of Future Consequences (CFCS) scale (OR 1.36, 95% CI 0.89 to 2.10), CFCS future subscale (OR 1.32, 95% CI 0.87 to 2.02) and CFCS immediate subscale (OR 1.35, 95% CI 0.88 to 2.07) ^68^ | - The odds of cessation at 4 weeks were significantly higher in those with than without post-school qualifications in analyses adjusted for all other variables including the Consideration of Future Consequences (CFCS) scale (OR 1.51, 95% CI 1.08 to 2.11), CFCS future subscale (OR 1.50, 95% CI 1.08 to 2.08) and CFCS immediate subscale (OR 1.10, 95% CI 1.10 to 2.14) ^68^ |
|  | Mental health UK Canada | No data | No data | No data |
|  | Secondary care UK Canada | No data | No data | No data |
| Age of leaving education (older) | General population UK |  | - The odds of cessation at the end of pregnancy were not significantly different among women who left education aged 19-21 years than those who left education aged ≥22 years in unadjusted (OR 0.69, 95% CI 0.43 to 1.10), mutually adjusted (OR 0.86, 95% CI 0.53 to 1.39) and mutually adjusted analyses including cigarette consumption (OR 0.98, 95% CI 0.58 to 1.67), and for those who left education aged 17-18 (versus those who left education aged ≥22) in mutually adjusted (OR 0.74, 95% CI 0.47 to 1.17) and mutually adjusted analysis including cigarette consumption (OR 0.84, 95% CI 0.51 to 1.40) ^76^ | - The odds of cessation at the end of pregnancy were significantly lower among women who left education aged ≤16 years than those who left education aged ≥22 years in unadjusted (OR 0.23, 95% CI 0.15 to 0.35), mutually adjusted (OR 0.47, 95% CI 0.30 to 0.74) and mutually adjusted analyses including cigarette consumption (OR 0.58, 95% CI 0.35 to 0.96), and for those who left education aged 17-18 (versus those who left education aged ≥22) in unadjusted analysis (OR 0.50, 95% CI 0.32 to 0.78) ^76^ |
|  | Mental health UK Canada | No data | No data | No data |
|  | Secondary care UK Canada | No data | No data | No data |
| Client pays for prescription (vs. exemption) | General population UK |  | - The odds of cessation at 4 weeks were not significantly different among those not eligible versus eligible for free prescriptions (OR 0.86, p=0.06) ^85^ | - The odds of cessation at 4 weeks were higher among those who paid for prescriptions than those with an exemption in adjusted analysis (OR 1.20, 95% CI 1.13 to 1.27) and an adjusted analysis of a sub-sample including only those with HSI data (OR 1.15, 95% CI 1.04 to 1.28) ^70^ - The odds of cessation at 4 weeks were lower among those who were exempt (OR 0.81, 95% CI 0.79 to 0.87) and those who did not disclose their prescription status (OR 0.80, 95% CI 0.77 to 0.83) than those who reported paying for prescriptions in adjusted analyses ^71^ |
|  | Mental health UK Canada | No data | No data | No data |
|  | Secondary care UK Canada | No data | No data | No data |
| Reading level (based on qualifications and usual newspaper) (high) | General population UK |  |  | - The odds of cessation at 6 months were higher among those with a high than low reading level (based in qualifications and usual newspaper) in univariate (OR 1.62, 95% CI 1.19 to 2.21) and multivariate (OR 1.85, 95% CI 1.04 to 3.27) analyses ^81^ |
|  | Mental health UK Canada | No data | No data | No data |
|  | Secondary care UK Canada | No data | No data | No data |
| Childhood NS-SEC socio-economic classification of mother (higher) | General population UK |  | - The odds of cessation in pregnancy was not significantly different in women whose mother worked in intermediate occupations than those whose mother worked in managerial/professional occupations in mutually adjusted analyses including cigarette consumption (OR 0.77, 95% CI 0.57 to 1.03) ^76^ | - The odds of cessation in pregnancy was significantly lower among women whose mother worked in routine and manual occupations, was long-term unemployed, and who didn’t know their mother’s occupation grade than those whose mother worked in a managerial/professional occupation in unadjusted analyses (OR 0.43, 95% CI 0.35 to 0.52; OR 0.27, 95% CI 0.20 to 0.36; OR 0.38, 95% CI 0.29 to 0.49), mutually adjusted analyses (OR 0.67, 95% CI 0.52 to 0.85; OR 0.50, 95% CI 0.36 to 0.71; OR 0.63, 95% CI 0.47 to 0.85) and mutually adjusted analyses including cigarette consumption (OR 0.70, 95% CI 0.54 to 0.91; OR 0.51, 95% CI 0.35 to 0.73; OR 0.69, 95% CI 0.50 to 0.93), and for those whose mother worked in intermediate occupations (vs. managerial/professional occupations) in unadjusted (OR 0.55, 95% CI 0.44 to 0.70) and mutually adjusted analyses (OR 0.74, 95% CI 0.56 to 0.97) ^76^ |
|  | Mental health UK Canada | No data | No data | No data |
|  | Secondary care UK Canada | No data | No data | No data |
| Employment status | General population UK |  | - The odds of smoking cessation at 4 weeks were not significantly different for those not in full-time employment than those who were (OR 1.12, p=0.19) in a univariate analysis ^85^ |  |
|  | Mental health UK Canada | No data | No data | No data |
|  | Secondary care UK Canada |  | - The odds of smoking cessation at around 9 weeks were not significantly different for those who were employed (OR 1.11, 95% CI 0.72 and 1.71) and unemployed (OR 0.82, 95% CI 0.52 to 1.30) than those who were retired in unadjusted analyses ^95^ - The odds of cessation at most-recent follow-up were not significantly different for those whose income source was a Canadian pension plan (OR 1.2, 95% CI 0.5 to 3.3) or earned income (OR 1.2, 95% CI 0.5 to 3.1) than those whose income source was disability or social assistance in an unadjusted analysis ^94^ |  |
| Social housing (vs. other housing) | General population UK | - The odds of cessation from the start of the most recent quit attempt to the time of the survey were significantly lower for those who lived in social housing than for those who lived in other housing in unadjusted (OR 0.53, 95% CI 0.42 to 0.66) and adjusted (OR 0.57, 95% CI 0.45 to 0.72) analyses ^80^ |  |  |
|  | Mental health UK Canada | No data | No data | No data |
|  | Secondary care UK Canada | No data | No data | No data |
| Marital status | General population UK | - The odds of cessation at 4 weeks were significantly higher for those who were single, divorced or separated than for those who were married or with a partner in univariate analysis (OR 1.43, p<0.001), analyses adjusted for all variables reaching significance in the univariate analyses (OR 1.33, p=0.003) and analysis adjusted for all variables reaching significance in the univariate analyses alongside choice of NRT product (OR 1.33, p=0.009) ^85^ | - The odds of cessation at 6 months were not significantly different for those who were married than for any unmarried status in multivariate analysis (OR 1.34, 95% CI 0.88 to 2.03) ^81^ | - The odds of cessation at 6 months were significantly higher for those who were married than any unmarried status in univariate analysis (OR 1.51, 95% CI 1.11 to 2.05) ^81^ |
|  | Mental health UK Canada | No data | No data | No data |
|  | Secondary care UK Canada |  |  | - The odds of cessation at around 9 weeks in those who were single were significantly lower than for those who were married in unadjusted analyses (OR 0.6, 95% CI 0.43 to 0.82) ^95^ |
| Cohabitation status (cohabiting) | General population UK |  | - The odds of cessation during pregnancy were not significantly different for lone mothers than for non-lone mothers in a mutually adjusted analysis (OR 1.00, 95% CI 0.81 to 1.23) and a mutually adjusted analysis including cigarette consumption (OR 1.00, 95% CI 0.80 to 1.25) ^76^ | - The odds of cessation during pregnancy were significantly lower for lone mothers than for non-lone mothers in an unadjusted analysis (OR 0.64, 95% CI 0.54 to 0.77 ^76^ |
|  | Mental health UK Canada | No data | No data | No data |
|  | Secondary care UK Canada | No data | No data | No data |
| Household smokers | General population UK |  | - The odds of cessation at 1 year were not significantly different for those who were the only household smoker than those who lived with (an)other smoker(s) in adjusted analyses of both smoking cessation services in England (OR 1.3, 95% CI 0.9 to 1.8) and one-to-one clinics in Glasgow (OR 3.0, 95% CI 0.8 to 11.2) ^77^ - The odds of cessation at 4 weeks were not significantly different for those with no other smokers in the household than for those with other smokers in the household (OR 0.96, p=0.78) ^85^ |  |
|  | Mental health UK Canada | No data | No data | No data |
|  | Secondary care UK Canada | No data | No data | No data |
| Social support to quit from family / friends (more) | General population UK |  | - The odds of cessation at 1 year were not significantly different for those who had support to quit than no support to quit in adjusted analyses of smoking cessation services in England (OR 1.6, 95% CI 0.8 to 3.4) and Glasgow one-to-one services (OR 1.0, 95% CI 0.3 to 3.8) ^77^ - The odds of cessation at 6 months were not significantly different as support from family and friends increased (1-5 scale) in an adjusted analysis (OR 1.03, 95% CI 0.89 to 1.20) ^81^ | - The odds of cessation at 6 months were significantly higher as support from family and friends increased (1-5 scale) in unadjusted analysis (OR 1.16, 95% CI 1.02 to 1.31) ^81^ |
|  | Mental health UK Canada |  | - The odds of cessation at 12 to 26 weeks were not significantly different for those who had social support for quitting than for those who did not (OR 4.8, 95% CI 1.0 to 22.3) in an unadjusted analysis ^88^ | - The odds of cessation at 12 to 26 weeks were significantly higher for those who had social support for quitting than for those who did not (OR 5.8, 95% CI 1.2 to 28.9) in an adjusted analysis ^88^ |
|  | Secondary care UK Canada | No data | No data | No data |
| **Health and healthcare setting** | | | | |
| Primary mental health diagnosis | General population UK | No data | No data | No data |
|  | Mental health UK Canada | - The odds of cessation at 8 to 28 weeks were significantly lower among those with anxiety disorder than for those with no disorder in unadjusted (OR 0.50, 95% CI 0.27 to 0.91) and adjusted analyses of the whole sample (OR 0.51, 95% CI 0.27 to 0.96) ^90^ | - The odds of cessation at 12 to 26 weeks were not significantly different for those with psychotic disorder compared with those with mood/anxiety disorder (OR 0.9, 95% CI 0.4 to 2.4) in a univariate analysis ^88^ - The odds of cessation at 8 to 28 weeks were not significantly different among those with mood disorder and those with psychotic disorder than for those with no disorder in unadjusted (OR 0.78, 95% CI 0.51 to 1.20; OR 0.65, 95% CI 0.31 to 1.39, respectively) and adjusted analyses of the whole sample (OR 0.79, 95% CI 0.49 to 1.25; OR 0.59, 95% CI 0.26 to 1.32, respectively) ^90^ - Among females, the odds of cessation at 8 to 26 weeks were not significantly different among those with mood disorder (OR 1.33, 95% CI 0.36 to 4.90), anxiety disorder (OR 1.19, 95% CI 0.27 to 5.14), and psychotic disorder (OR 7.05, 95% CI 0.68 to 72.60) compared with those with no disorder in unadjusted analyses ^89^ - The odds of cessation at end of treatment were not significantly different for those with schizophrenia compared with those without schizophrenia (OR 0.82, 95% CI 0.34 to 1.99) in adjusted analysis of the whole sample ^91^ |  |
|  | Secondary care UK Canada | No data | No data | No data |
| Hospital Anxiety and Depression Scale (HADS) anxiety score | General population UK | No data | No data | No data |
|  | Mental health UK Canada | No data | No data | No data |
|  | Secondary care UK Canada |  | - The odds of cessation at around 9 weeks were not significantly different according to HADS anxiety score (continuous) in an unadjusted analysis (OR 1.03, 95% CI 0.98 to 1.08) ^95^ |  |
| HADS depression score | General population UK | No data | No data | No data |
|  | Mental health UK Canada | No data | No data | No data |
|  | Secondary care UK Canada |  | - The odds of cessation at around 9 weeks were not significantly different according to HADS depression score (continuous) in an unadjusted analysis (OR 0.95, 95% CI 0.90 to 1.00) ^95^ |  |
| Patient Health Questionnaire-9 (PHQ-9) depression score | General population UK | No data | No data | No data |
|  | Mental health UK Canada |  | - The odds of cessation at 12 to 26 weeks were not significantly impacted by PHQ-9 score (continuous) (OR 1.0, 95% CI 1.0 to 1.1) in a univariate analysis ^88^ - The odds of cessation at 3 months were not significantly different for those with mild depression and those with moderate to severe depression compared with those with none to minimal depression in unadjusted (OR 0.89, 95% CI 0.62 to 1.27; OR 0.83, 95% CI 0.58 to 1.20, respectively) and adjusted (OR 1.15, 95% CI 0.75 to 1.77; OR 1.05, 95% CI 0.67 to 1.66, respectively) analyses ^93^ - The odds of cessation at 6 months were not significantly different for those with mild depression and those with moderate to severe depression compared with those with none to minimal depression in unadjusted (OR 1.01, 95% CI 0.66 to 1.52; OR 0.93, 95% CI 0.61 to 1.43, respectively) and adjusted (OR 1.22, 95% CI 0.77 to 1.94; OR 1.15, 95% CI 0.70 to 1.89, respectively) analyses ^93^ |  |
|  | Secondary care UK Canada | No data | No data | No data |
| History of depression | General population UK | No data | No data | No data |
|  | Mental health UK Canada | - The odds of self-reported abstinence at 6 months were significantly lower among those with current or recent depression (OR 0.71, 95% CI 0.53 to 0.96) and recurrent depression (OR 0.38, 95% CI 0.20 to 0.72) than those with no history of depression in an unadjusted and adjusted analyses (OR 0.74, 95% CI 0.51 to 1.07; OR 0.45, 95% CI 0.21 to 0.96, respectively) ^92^ - The odds of 30-day point-prevalent abstinence at 6 months were significantly lower among those with recurrent depression than those with no history of depression in unadjusted (OR 0.62, 95% CI 0.46 to 0.83) and adjusted (OR 0.65, 95% CI 0.45 to 0.94) analyses ^92^ | - The odds of self-reported abstinence at 6 months were not significantly different for those with past depression (OR 0.89, 95% CI 0.66 to 1.21) than those with no history of depression in an unadjusted analysis, and for those with past depression (OR 1.09, 95% CI 0.76 to 1.56) and current / recent depression (OR 0.74, 95% CI 0.51 to 1.07) in an adjusted analysis ^92^ - The odds of 30-day point-prevalent at 6 months were not significantly different for those with past depression (OR 0.96, 95% CI 0.80 to 1.16) than those with no history of depression in an unadjusted analysis, and for those with past depression (OR 0.96, 95% CI 0.77 to 1.19) and current / recent depression (OR 0.82, 95% CI 0.67 to 1.02) in an adjusted analysis ^92^ |  |
|  | Secondary care UK Canada | No data | No data | No data |
| History of psychiatric disorder | General population UK | No data | No data | No data |
|  | Mental health UK Canada | No data | No data | No data |
|  | Secondary care UK Canada |  | - The odds of cessation at most recent follow-up were not significantly different for those with history of a psychiatric disorder (OR 0.3, 95% CI 0.1 to 1.0), history of a substance use disorder (OR 0.9, 95% CI 0.3 to 2.4) and both (OR 0.5, 95% CI 0.2 to 1.3) compared with those with no history of either psychiatric nor substance use disorder, in an unadjusted analysis ^94^ |  |
| History of substance abuse | General population UK | No data | No data | No data |
|  | Mental health UK Canada | - The odds of cessation at 8 to 28 weeks were significantly lower for those with a history of opiate abuse (OR 0.43, 95% CI 0.19 to 0.95) than for those with a history of alcohol abuse in an adjusted analysis of a subsample with co-occurring disorders ^90^ - The odds of cessation at 8 to 26 weeks were significantly lower for those with a history of alcohol abuse (OR 0.32, 95% CI 0.11 to 0.94), opiate abuse (OR 0.15, 95% CI 0.04 to 0.60), and marijuana abuse (OR 0.17, 95% CI 0.05 to 0.61) than for those with no history of substance abuse in an adjusted analysis of the whole sample, and for those with a history of alcohol abuse (OR 0.19, 95% CI 0.04 to 0.82), opiate abuse (OR 0.09, 95% CI 0.01 to 0.67) and marijuana abuse (OR 0.14, 95% CI 0.02 to 0.85) than those with no history of substance abuse in an adjusted analysis of a subsample of males ^89^ | - The odds of cessation at 12 to 26 weeks were not significantly different for those with a history of substance abuse than those with no history of substance abuse (OR 0.3, 95% CI 0.5 to 3.4) in a univariate analysis ^88^ - The odds of cessation at 8 to 28 weeks were not significantly different for those with a history of alcohol abuse (OR 0.88, 95% CI 0.50 to 1.53), opiate abuse (OR 0.62, 95% CI 0.31 to 1.24), cocaine abuse (OR 1.11, 95% CI 0.62 to 1.97), marijuana abuse (OR 0.79, 95% CI 0.39 to 1.62) and methamphetamine abuse (OR 0.99, 95% CI 0.37 to 2.65) than for those with no history of substance abuse in an unadjusted analysis of the whole sample, and for those with a history of cocaine abuse (OR 1.29, 95% CI 0.73 to 2.30), marijuana abuse (OR 0.87, 95% CI 0.42 to 1.77) and methamphetamine abuse (OR 1.36, 95% CI 0.46 to 4.05) than those with a history of alcohol abuse in an adjusted analysis of a subsample with co-occurring disorders ^90^ - The odds of cessation at 8 to 26 weeks were not significantly different for those with a history of cocaine abuse and those with a history of methamphetamine abuse than for than for those with no history of substance abuse in an adjusted analysis of the whole sample (OR 0.81, 95% CI 0.29 to 2.33; OR 0.19, 95% CI 0.03 to 1.28, respectively), and in an adjusted analysis of a subsample of males (OR 0.49, 95% CI 0.11 to 2.16; OR 0.10, 95% CI 0.00 to 2.70, respectively) ^89^ |  |
|  | Secondary care UK Canada | No data | No data |  |
| Co-occurring disorder | General population UK | No data | No data |  |
|  | Mental health UK Canada | No data | No data |  |
|  | Secondary care UK Canada |  | - The odds of cessation at 8 to 28 weeks were not significantly different in those with a substance use disorder only (OR 1.00, 95% CI 0.36 to 2.80), a psychiatric disorder only (OR 0.81, 95% CI 0.27 to 2.41) and co-occurring (psychiatric and substance use) disorder (OR 0.96, 95% CI 0.26 to 1.84) than those with neither a psychiatric nor substance use disorder, in an unadjusted analysis ^94^ |  |
| Cardiovascular risk | General population UK | No data | No data | No data |
|  | Mental health UK Canada | No data | No data | No data |
|  | Secondary care UK Canada |  | - The odds of cessation at around 9 weeks were not significantly different for those with moderate than high cardiovascular risk (OR 1.15, 95% CI 0.75 to 1.78) ^95^ | - The odds of cessation at around 9 weeks were significantly higher for those with low than high cardiovascular risk (OR 1.71, 95% CI 1.12 to 2.62) in an unadjusted analysis ^95^ |
| Comorbidities (more) | General population UK | No data | No data | No data |
|  | Mental health UK Canada | No data | No data | No data |
|  | Secondary care UK Canada | - The odds of cessation at around 9 weeks were significantly worse as comorbidities (continuous) increased (OR 0.88, 95% CI 0.80 to 0.96) in an unadjusted analysis ^95^ | - The odds of cessation at most recent follow-up were not significantly different as the number of medical comorbidities (continuous) increased (OR 1.0, 95% CI 0.7 to 1.4) in an unadjusted analysis ^94^ |  |
| Referral source | General population UK | No data | No data | No data |
|  | Mental health UK Canada | No data | No data | No data |
|  | Secondary care UK Canada |  | - The odds of cessation were not significantly different for those who were referred through Cardiology (OR 1.7, 95% CI 0.4 to 6.8) or Respirology (OR 1.5, 95% CI 0.3 to 8.8) than for those who were referred through another source in an unadjusted analysis ^94^ |  |
| **Tobacco smoking variables** | | | | |
| Daily cigarette consumption | General population UK | - The odds of cessation at 6 months significantly decreased as daily cigarette consumption (continuous) increased (OR 0.79, 95% CI 0.70 to 0.88) in an unadjusted analysis ^74^ | - The odds of cessation at 6 months were not significantly impacted by daily cigarette consumption (continuous) in an adjusted analysis (OR 0.82, 95% CI 0.65 to 1.03) ^74^ |  |
|  | Mental health UK Canada |  | - The odds of cessation at 12 to 26 weeks were not significantly impacted by average number of cigarettes per day (continuous) in a univariate analysis (OR 1.0, 95% CI 0.9 to 1.0) ^88^ - The odds of cessation at 8 to 28 weeks were not significantly impacted by average number of cigarettes per day (continuous) in a univariate analysis (OR 0.99, 95% CI 0.97 to 1.00) ^90^ |  |
|  | Secondary care UK Canada |  | - The odds of cessation at most recent follow-up were not significantly impacted by average number of cigarettes per day (continuous) in an unadjusted analysis (OR 1.0, 95% CI 0.9 to 1.0) ^94^ |  |
| Daily cigarette consumption before pregnancy | General population UK | - The odds of cessation in pregnancy were significantly lower among those who smoked 10-19 and ≥20 cigarettes per day than among those who smoked 1-9 cigarettes per day in unadjusted analyses (OR 0.29, 95% CI 0.25 to 0.34; OR 0.15, 95% CI 0.12 to 0.18, respectively) and mutually adjusted analyses including cigarette consumption (OR 0.31, 95% CI 0.27 to 0.37; OR 0.17, 95% CI 0.14 to 0.21, respectively) ^76^ |  |  |
|  | Mental health UK Canada | No data | No data | No data |
|  | Secondary care UK Canada | No data | No data | No data |
| CO level at baseline | General population UK | No data | No data | No data |
|  | Mental health UK Canada | - The odds of cessation at 8 to 28 weeks were significantly lower as baseline CO level (continuous) increased, in unadjusted analyses (OR 0.95, 95% CI 0.92 to 0.98, adjusted analyses of the whole sample (OR 0.93, 95% CI 0.88 to 0.98) and a sub-sample of those with co-occurring psychiatric and substance use disorders (OR 0.96, 95% CI 0.92 to 1.00) ^89^ | - The odds of cessation at 8 to 28 weeks were not significantly different by baseline CO level (continuous), in unadjusted analyses (OR 0.99, 95% CI 0.97 to 1.00), adjusted analyses of the whole sample (OR 0.99, 95% CI 0.98 to 1.01) and a sub-sample of those with co-occurring psychiatric and substance use disorders (OR 0.98, 95% CI 0.96 to 1.00) ^90^ |  |
|  | Secondary care UK Canada |  | - The odds of cessation at most recent follow-up were not significantly different by baseline expired CO level (continuous) in an unadjusted analysis (OR 0.9, 95% CI 0.9 to 1.0) nor an adjusted analysis (OR 0.9, 95% CI 0.9 to 1.0) ^94^ |  |
| Age started smoking | General population UK | - The odds of cessation at 6 months were significantly greater for those who started smoking aged >16 years than those who started smoking <14 years (OR 2.00, 95% CI 1.14 to 3.50) in a univariate analysis ^81^ | - The odds of cessation at 6 months were not significantly different for those who started smoking aged 14-16 years than those who started smoking aged <14 years in an unadjusted analysis (OR 1.65, 95% CI 0.93 to 2.91), and for those who started smoking aged 14-16 years (OR 1.25, 95% CI 0.67 to 2.35) and those who started smoking aged >16 years (OR 1.36, 95% CI 0.71 to 2.61) than those who started smoking aged <14 years in a multivariate analysis ^81^ |  |
|  | Mental health UK Canada |  | - The odds of cessation at 12 to 26 weeks were not significantly impacted by age of smoking initiation (continuous) in a univariate analysis (OR 1.0, 95% CI 1.0 to 1.1) ^88^   The odds of cessation at 8 to 28 weeks were not significantly impacted by age of smoking initiation (continuous) in an unadjusted analysis (OR 1.01, 95% CI 0.98 to 1.04) ^90^ |  |
|  | Secondary care UK Canada |  | - The odds of cessation at most recent follow-up were not significantly impacted by age of smoking initiation (continuous) an unadjusted analysis (OR 1.0, 95% CI 1.0 to 1.1) ^94^ |  |
| Heaviness of smoking index (HIS) | General population UK | - The odds of cessation at 4 weeks were significantly lower as HSI score increased (per unit increase) in an adjusted analysis of a sub-sample of only those with data on HSI score (OR 0.88, 95% CI 0.85 to 0.90) ^70^ - The odds of cessation at 6 months were significantly lower as HSI score (continuous) increased, in univariate (OR 0.88, 95% CI 0.82 to 0.93), multivariate (OR 0.84, 95% CI 0.78 to 0.89) and multiple imputation (OR 0.85, 95% CI 0.79 to 0.92) analyses, and for those in HSI category 5 (univariate OR 0.60, 95% CI 0.38 to 0.97; multivariate OR 0.52, 95% CI 0.31 to 0.86; multiple imputation OR 0.54, 95% CI 0.31 to 0.95) and category 6 (univariate OR 0.37, 95% CI 0.20 to 0.69; multivariate OR 0.26, 95% CI 0.13 to 0.51; multiple imputation OR 0.25, 95% CI 0.13 to 0.48) than those in category 0 ^86^ - The odds of cessation at 6 months were significantly lower for those with medium (OR 0.59, 95% CI 0.38 to 0.90) and high (OR 0.42, 95% CI 0.29 to 0.62) than low nicotine dependence index in a univariate analysis, and for those with high (OR 0.58, 95% CI 0.34 to 0.98) than low nicotine dependence in a multivariate analysis ^81^ | - The odds of cessation were not significantly different for those in HSI category 1 (univariate OR 0.97, 95% CI 0.60 to 1.57; multivariate OR 1.05, 95% CI 0.63 to 1.74; multiple imputation OR 1.10, 95% CI 0.63 to 1.95), category 2 (univariate OR 1.01, 95% CI 0.65 to 1.58; multivariate OR 0.98, 95% CI 0.61 to 1.58; multiple imputation OR 0.95, 95% CI 0.55 to 1.66), category 3 (univariate OR 0.76, 95% CI 0.49 to 1.17; multivariate OR 0.78, 95% CI 0.49 to 1.23; multiple imputation OR 0.79, 95% CI 0.48 to 1.32), or category 4 (univariate OR 0.76, 95% CI 0.49 to 1.17; multivariate OR 0.70, 95% CI 0.44 to 1.11; multiple imputation OR 0.83, 95% CI 0.50 to 1.39), than those in HSI category 0 ^86^ - The odds of cessation at 6 months were not significantly different for those with medium than low nicotine dependence index (OR 0.65, 95% CI 0.38 to 1.11) in a multivariate analysis ^81^ |  |
|  | Mental health UK Canada | No data | No data | No data |
|  | Secondary care UK Canada | No data | No data | No data |
| Fagerstrom Test for Nicotine Dependence (FTND) | General population UK | - The odds of cessation at 4 weeks were significantly lower as FTND (continuous) increased (OR 0.95, 95% CI 0.94 to 0.97) in an adjusted analysis ^87^ - The odds of cessation at 4 weeks were significantly lower as baseline FTND (continuous) increased, in a univariate analysis (OR 0.93, p<0.001), multivariate analysis adjusted for all variables reaching significance in the univariate analyses (OR 0.94, p=0.006), and multivariate analysis adjusted for all variables reaching significance in the univariate analyses alongside choice of NRT product (OR 0.95, p=0.95) ^85^ - The odds of cessation at 4 weeks were significantly lower as baseline FTND score (continuous) increased, in analyses adjusted for all other variables including the Consideration of Future Consequences (CFCS) scale (OR 0.89, 95% CI 0.82 to 0.96), CFCS future subscale (OR 0.89, 95% CI 0.82 to 0.96) and CFCS immediate subscale (OR 0.89, 95% CI 0.82 to 0.96) ^68^ - The odds of cessation at 6 months were significantly lower as baseline FTND score (continuous) increased, in analyses adjusted for all other variables including the Consideration of Future Consequences (CFCS) scale (OR 0.88, 95% CI 0.80 to 0.97), CFCS future subscale (OR 0.88, 95% CI 0.80 to 0.97) and CFCS immediate subscale (OR 0.88, 95% CI 0.79 to 0.97) ^68^ |  |  |
|  | Mental health UK Canada | - The odds of cessation at 12 to 26 weeks were significantly lower as baseline FTND (continuous) increased (OR 0.7, 95% CI 0.6 to 0.9) in a multivariate analysis ^88^ - The odds of cessation at 8 to 28 weeks were significantly lower as baseline FTND (continuous) increased, in unadjusted analyses (OR 0.89, 95% CI 0.82 to 0.97, adjusted analyses of the whole sample (OR 0.90, 95% CI 0.82 to 0.98) and a sub-sample of those with co-occurring psychiatric and substance use disorders (OR 0.85, 95% CI 0.75 to 0.97) ^90^ | - The odds of cessation at 12 to 26 weeks were not significantly impacted by FTND at baseline (continuous) in a univariate analysis (OR 0.8, 95% CI 0.6 to 1.0) ^88^ - The odds of cessation at 8 to 26 were not significantly impacted by FTND score (continuous) in an adjusted analysis of the total sample (OR 0.96, 95% CI 0.79 to 1.16) nor an adjusted analysis of a male-only sub-sample (OR 0.90, 95% CI 0.70 to 1.15) ^89^ - The odds of cessation at end of treatment were not significantly different by baseline FTND score (continuous) in an adjusted analysis (OR 1.27, 95% CI 0.98 to 1.64) ^91^ |  |
|  | Secondary care UK Canada |  | - The odds of cessation at most recent follow-up were not significantly different by FTND score (continuous) in an unadjusted analysis (OR 0.8, 95% CI 0.7 to 1.0) nor an adjusted analysis (OR 0.9, 95% CI 0.7 to 1.1) ^94^ |  |
| Time to first daily smoke | General population UK |  | - The odds of cessation at 1 year were not significantly different for those whose first daily smoke was more than 5 minutes of waking than within 5 minutes (OR 1.2, 95% CI 0.9 to 1.8) in smoking cessation services in England, nor for those whose first daily smoke was more than 1 hour of waking than within 1 hour (OR 3.4, 95% CI 0.7 to 15.9) in one-to-one clinics in Glasgow, in adjusted analyses ^77^ |  |
|  | Mental health UK Canada | No data | No data | No data |
|  | Secondary care UK Canada | No data | No data | No data |
| Strength of urges to smoke | General population UK | - The odds of cessation at 6 months were significantly lower for those who reported strong urges to smoke than none in unadjusted (OR 0.55, 95% CI 0.36 to 0.83) and adjusted analyses (OR 0.59, 95% CI 0.37 to 0.93) ^74^ - The odds of cessation at 6 months were significantly lower as strength of urges to smoke (continuous) increased (OR 0.77, 95% CI 0.66 to 0.89) in adjusted analysis ^78^ - The odds of cessation at 4 weeks to 6 months were significantly lower as strength of urges to smoke increased (per point increase) in a univariate analysis (OR 0.70, 95% CI 0.57 to 0.87), analysis adjusted for strength of urges/enjoyment (OR 0.70, 95% CI 0.57 to 0.87) and an analysis adjusted for adjusted for strength of urges/enjoyment plus gender, age and social grade (OR 0.72, 95% CI 0.58 to 0.89) ^73^ - The odds of cessation at 6 months were significantly lower as strength of urges to smoke increased (continuous) in a multiple imputation analysis (OR 0.94, 95% CI 0.80 to 0.99) ^86^ | - The odds of cessation at 6 months were not significantly different for those with slight (unadjusted OR 1.32, 95% CI 0.87 to 2.03; adjusted OR 1.27, 95% CI 0.81 to 2.01), moderate (unadjusted OR 0.74, 95% CI 0.51 to 1.09; adjusted OR 0.80, 95% CI 0.54 to 1.21), very strong (unadjusted OR 0.60, 95% CI 0.34 to 1.02; adjusted OR 0.71, 95% CI 0.39 to 1.28) and extremely strong urges to smoke (unadjusted OR 0.65, 95% CI 0.28 to 1.38; adjusted OR 0.88, 95% CI 0.36 to 2.04) than those who reported none, in unadjusted and adjusted analyses ^74^ - The odds of cessation at 6 months were not significantly impacted by strength of urges to smoke (continuous) in univariate or multivariate analyses, and were not significantly different for those in strength of urges to smoke item category 2, 3, 4, 5 or 6 than category 1 (lowest strength of urges to smoke) in univariate, multivariate and multiple imputation analyses ^86^ |  |
|  | Mental health UK Canada | No data | No data | No data |
|  | Secondary care UK Canada | No data | No data | No data |
| Frequency of urges to smoke | General population UK | - The odds of cessation at 6 months were significantly lower as frequency of urges to smoke increased (continuous) in a univariate analysis (OR 0.89, 95% CI 0.81 to 0.97), multivariate analysis (OR 0.88, 95% CI 0.81 to 0.97), and multiple imputation analysis (OR 0.89, 95% CI 0.80 to 0.99) ^86^ | - The odds of cessation at 6 months were not significantly different for those in strength of urges to smoke item category 2, 3, 4, 5 or 6 than category 1 (lowest strength of urges to smoke) in univariate, multivariate and multiple imputation analyses ^86^ |  |
|  | Mental health UK Canada | No data | No data | No data |
|  | Secondary care UK Canada | No data | No data | No data |
| Most difficult situation not to smoke | General population UK | - The odds of cessation at 6 months were significantly higher in those whose most difficult situation not to smoke was when feeling the urge to smoke (OR 1.72, 95% CI 1.16 to 2.55) and when relaxing (OR 1.95, 95% CI 1.08 to 3.51) than for those whose most difficult situation not to smoke was when anxious or stressed in a univariate analysis, and for those whose most difficult situation not to smoke was when feeling the urge to smoke (OR 1.65, 95% CI 1.06 to 2.58) than when anxious or stressed in a multivariate analysis ^81^ | - The odds of cessation at 6 months were not significantly different between those whose most difficult situation not to smoke was when socialising (OR 1.46, 95% CI 0.88 to 2.42), first thing in the morning (OR 0.79, 95% CI 0.47 to 1.31), when angry or frustrated (OR 1.06, 95% CI 0.57 to 2.00), and for any other reason (OR 1.34, 95% CI 0.39 to 4.64), than when anxious or stressed in a univariate analysis, and when socialising (OR 1.24, 95% CI 0.68 to 2.24), first thing in the morning (OR 1.03, 95% CI 0.58 to 1.82), when angry or frustrated (OR 1.11, 95% CI 0.56 to 2.17), when relaxing (OR 1.68, 95% CI 0.83 to 3.41) or another reason (OR 0.59, 95% CI 0.14 to 2.52) than when anxious or stressed, in a multivariate analysis ^81^ |  |
|  | Mental health UK Canada | No data | No data | No data |
|  | Secondary care UK Canada | No data | No data | No data |
| Enjoyment of smoking | General population UK |  | - The odds of cessation at 4 weeks to 6 months were not significantly impacted by enjoyment of smoking (per point increase) in a univariate analysis (OR 0.99, 95% CI 0.77 to 1.27), analysis adjusted for strength of urges/enjoyment (OR 1.04, 95% CI 0.80 to 1.34) and an analysis adjusted for adjusted for strength of urges/enjoyment plus gender, age and social grade (OR 1.01, 95% CI 0.77 to 1.31) ^73^ |  |
|  | Mental health UK Canada | No data | No data | No data |
|  | Secondary care UK Canada | No data | No data | No data |
| Determination to quit | General population UK | - The odds of cessation at 1 year were significantly higher for those who were extremely determined than those who were not at all/quite/very determined to quit in adjusted analyses of smoking cessation services in England ^77^ | - The odds of cessation at 1 year were not significantly different for those who were extremely determined than those who were not at all/quite/very determined to quit in adjusted analyses of one-to-one clinics in Glasgow (OR 2.4, 95% CI 0.8 to 7.5) ^77^ |  |
|  | Mental health UK Canada | No data | No data | No data |
|  | Secondary care UK Canada | No data | No data | No data |
| Motivation to stop smoking | General population UK | - The odds of cessation at 1 year were significantly higher for those who expressed motivation to stop in the next three months than those with no motivation to stop in an adjusted analysis (OR 2.54, 95% CI 1.81 to 3.56) ^78^ |  |  |
|  | Mental health UK Canada | No data | No data | No data |
|  | Secondary care UK Canada | No data | No data | No data |
| Importance of quitting | General population UK | No data | No data | No data |
|  | Mental health UK Canada |  | - The odds of cessation at 12 to 26 weeks were not significantly different by the importance of quitting (continuous) in a univariate analysis (OR 1.1, 95% CI 0.8 to 1.5) ^88^ - The odds of cessation at 8 to 28 weeks were not significantly different by the importance of quitting (continuous) in an unadjusted analysis (OR 1.04, 95% CI 0.90 to 1.20) ^90^ |  |
|  | Secondary care UK Canada |  | - The odds of cessation at most recent follow-up were not significantly different by the importance of quitting (continuous) in an unadjusted analysis (OR 1.1, 95% CI 0.9 to 1.3) ^94^ |  |
| Confidence in quitting | General population UK | No data | No data | No data |
|  | Mental health UK Canada |  | - The odds of cessation at 12 to 26 weeks were not significantly different by confidence in quitting (continuous) in a univariate analysis (OR 1.0, 95% CI 0.8 to 1.2) ^88^ - The odds of cessation at 8 to 28 weeks were not significantly different by confidence in quitting (continuous) in an unadjusted analysis (OR 1.03, 95% CI 0.96 to 1.11) ^90^ |  |
|  | Secondary care UK Canada |  | - The odds of cessation at most recent follow-up were not significantly different by confidence in quitting (continuous) in an unadjusted analysis (OR 1.2, 95% CI 1.0 to 1.4) or an adjusted analysis (OR 1.1, 95% CI 0.9 to 1.3) ^94^ |  |
| Readiness to quit | General population UK | No data | No data | No data |
|  | Mental health UK Canada |  | - The odds of cessation at 12 to 26 weeks were not significantly different by readiness to quit (continuous) in a univariate analysis (OR 1.1, 95% CI 0.9 to 1.3) ^88^ |  |
|  | Secondary care UK Canada | No data | No data | No data |
| Stage of readiness to quit (Perspectives of Change Model) | General population UK |  | - The odds of cessation at 6 months were not significantly different for those who were in the engaged (OR 1.24, 95% CI 0.841 to 1.92) or disengaged stage of readiness and planning to quit within the next 6 months (OR 0.88, 95% CI 0.59 to 1.31) than those in the disengaged stage of readiness and not planning to quit within the next 6 months (of the Perspectives of Change Model) in a univariate analysis, and for those who in the committed stage of readiness (OR 1.91, 95% CI 0.91 to 3.99), engaged (OR 1.69, 95% CI 0.98 to 2.91) or disengaged stage of readiness and planning to quit within the next 6 months (OR 0.98, 95% CI 0.61 to 1.57) than those in the disengaged stage of readiness and not planning to quit within the next 6 months (of the Perspectives of Change Model), in a multivariate analysis ^81^ | - The odds of cessation at 6 months were significantly higher for those who were in the committed stage of readiness than those in the disengaged stage of readiness and not planning to quit within the next 6 months (of the Perspectives of Change Model) in a univariate analysis (OR 1.97, 95% CI 1.10 to 3.52) ^81^ |
|  | Mental health UK Canada | No data | No data | No data |
|  | Secondary care UK Canada | No data | No data | No data |
| Stage of readiness to quit (individual items) | General population UK | - The odds of cessation at 6 months were significantly lower for those who indicated greater agreement with the item ‘Think of yourself as addicted to smoking’ (1-5 scale) in a univariate analysis (OR 0.77, 95% CI 0.88 to 1.23) ^81^ | - The odds of cessation at 6 months were not significantly different for different responses (on a 1-5 scale) to the individual items ‘How much do you want to quit?’, 'How determined are you to quit for good?', 'Smoking is part of who I am', and 'I can see myself as a non-smoker' in both univariate and multivariate analyses, and for the item ‘Think of yourself as a addicted to smoking’ in a multivariate analysis ^81^ |  |
|  | Mental health UK Canada | No data | No data | No data |
|  | Secondary care UK Canada | No data | No data | No data |
| Stage of Change (Transtheoretical Model) | General population UK | No data | No data | No data |
|  | Mental health UK Canada |  | - The odds of cessation at 12 to 26 weeks were not significantly different for those in the preparation or action stage than those in the precontemplation or contemplation stage of the Transtheoretical Model in a univariate analysis (OR 2.4, 95% CI 0.8 to 6.7) ^88^ |  |
|  | Secondary care UK Canada |  | - The odds of cessation at most recent follow-up were not significantly different for those in the preparation or action stage of change than for those in the precontemplation or contemplation stage of the Transtheoretical Model, in an adjusted analysis (OR 3.3, 95% CI 1.0 to 11.3) ^94^ | - The odds of cessation at most recent follow-up were significantly higher for those in the preparation or action stage of change than for those in the precontemplation or contemplation stage of the Transtheoretical Model, in an unadjusted analysis (OR 4.7, 95% CI 1.7 to 13.3) ^94^ |
| Reason for quitting | General population UK |  | - The odds of cessation at most recent follow-up were not significantly different for those whose reason for quitting was to gain control (unadjusted OR 1.06, 95% CI 0.72 to 1.56; adjusted OR 1.15, 95% CI 0.69 to 1.91), to save money (unadjusted OR 0.78, 95% CI 0.42 to 1.43; adjusted OR 0.95, 95% CI 0.46 to 1.97), pressure from others (unadjusted OR 1.53, 95% CI 0.85 to 2.76; adjusted OR 1.03, 95% CI 0.45 to 2.34), or another reason (unadjusted OR 0.86, 95% CI 0.39 to 1.89; adjusted OR 0.35, 95% CI 0.11 to 1.14), than for those whose reason for quitting was concern about illness ^81^ |  |
|  | Mental health UK Canada | No data | No data | No data |
|  | Secondary care UK Canada | No data | No data | No data |
| Main advantage of quitting | General population UK |  | - The odds of cessation at 6 months were not significantly different for those for whom the main advantage of quitting was a sense of achievement, being more energetic and alert, feeling calm and content, being able to taste food better, and any other reason, than for those for whom the main advantage of quitting was enjoying good health, in univariate and multivariate analyses ^81^ |  |
|  | Mental health UK Canada | No data | No data | No data |
|  | Secondary care UK Canada | No data | No data | No data |
| Main disadvantage of quitting | General population UK |  | - The odds of cessation at 6 months were not significantly different among those for whom the main disadvantage of quitting was loss of concentration (OR 0.33, 95% CI 0.08 to 1.40), gaining weight (OR 1.28, 95% CI 0.94 to 1.74), and feeling dull and bored (OR 1.48, 95% CI 0.86 to 2.54) than for those for whom the main disadvantage of quitting was increased tension, in a univariate analysis, and for those for whom the main disadvantage of quitting was loss of concentration (OR 0.28, 95% CI 0.06 to 1.28), gaining weight (OR 1.02, 95% CI 0.70 to 1.48), feeling dull and bored (OR 1.04, 95% CI 0.56 to 1.95), and any other disadvantage (OR 1.22, 95% CI 0.68 to 2.17) than for those for whom the main disadvantage of quitting was increased tension, in a multivariate analysis ^81^ | - The odds of cessation at 6 months were significantly higher among those for whom the main disadvantage of quitting was any other disadvantage (other than loss of concentration, gaining weight, and feeling dull and bored) than for those for whom the main disadvantage of quitting was experiencing increased tension (OR 1.91, 95% CI 1.12 to 3.26) in a univariate analysis ^81^ |
|  | Mental health UK Canada | No data | No data | No data |
|  | Secondary care UK Canada | No data | No data | No data |
| Past serious quit attempt | General population UK |  | - The odds of cessation at 4 weeks were not significantly different for those with no past serious quit attempt than for those with a at least one past serious quit attempt in a univariate analysis (OR 1.22, p=0.06) ^85^ |  |
|  | Mental health UK Canada | No data | No data | No data |
|  | Secondary care UK Canada | No data | No data | No data |
| Smoking cessation treatment episode (later vs. first) | General population UK |  | - The odds of cessation at 4 weeks were not significantly different for those undergoing a later cessation treatment episode than for those undergoing their first treatment episode (OR 0.99, 95% CI 0.90 to 1.08) in an adjusted analysis ^69^ |  |
|  | Mental health UK Canada | No data | No data | No data |
|  | Secondary care UK Canada | No data | No data | No data |
| Previously quit smoking for 3 months or more | General population UK |  |  | - The odds of cessation at 6 months were significantly higher for those who had than those who had not previously quit smoking for 3 months or more in univariate (OR 1.81, 95% CI 1.28 to 2.58) and multivariate (OR 1.58, 95% CI 1.05 to 2.39) analyses ^81^ |
|  | Mental health UK Canada | No data | No data | No data |
|  | Secondary care UK Canada | No data | No data | No data |
| Number of past cessation attempts | General population UK | No data | No data | No data |
|  | Mental health UK Canada |  | - The odds of cessation at 12 to 26 weeks were not significantly different for who had made more than 5 quit attempts than for those who had made 5 attempts or fewer, in a univariate analysis (OR 0.9, 95% CI 0.3 to 2.8) ^88^ |  |
|  | Secondary care UK Canada | No data | No data | No data |
| Number of quit attempts past 6 months | General population UK |  |  | - The odds of cessation at 6 months were significantly higher for those who had made two (unadjusted OR 0.43, 95% CI 0.32 to 0.57; adjusted OR 0.45, 95% CI 0.33 to 0.62) and those who had made three or more (unadjusted OR 0.30, 95% CI 0.18 to 0.47; adjusted OR 0.32, 95% CI 0.19 to 0.51) than for those who had made one quit attempt in the past 6 months ^74^ |
|  | Mental health UK Canada | No data | No data | No data |
|  | Secondary care UK Canada | No data | No data | No data |
| Quit for how long at last quit attempt | General population UK | No data | No data | No data |
|  | Mental health UK Canada |  | - The odds of cessation at 8 to 28 weeks did not significantly differ by length of cessation at the last quit attempt (continuous) in an adjusted analysis of a sub-sample of those with a psychiatric disorder (OR 1.48, 95% CI 0.99 to 2.22) ^90^ | - The odds of cessation at 8 to 28 weeks were significantly higher with greater length of cessation at the last quit attempt (continuous) in an unadjusted analysis (OR 1.13, 95% CI 1.02 to 1.27) and an adjusted analysis of the whole sample (OR 1.14, 95% CI 1.01 to 1.28) ^90^ |
|  | Secondary care UK Canada |  | - The odds of cessation at most recent follow-up were not significantly different for those who had quit for 1 week to less than 1 month (OR 1.7, 95% CI 0.4 to 7.0), and more than 1 year (OR 2.0, 95% CI 0.6 to 6.8) at the last attempt, than for those who had quit for less than 1 week at the last attempt, in an unadjusted analysis, and for those who had quit for 1 week to less than 1 month (OR 1.5, 95% CI 0.3 to 9.1), 7 months to 1 year (OR 2.6, 95% CI 0.6 to 11.5), and more than 1 year (OR 1.8, 95% CI 0.4 to 7.9) at the last attempt than for those who had quit for less than 1 week at the last attempt, in an adjusted analysis ^94^ | - The odds of cessation at most recent follow-up were significantly higher for those who had quit for 7 months to 1 year at the last attempt (OR 3.6, 95% CI 1.1 to 11.5) than for those who had quit for less than 1 week at the last attempt, in an unadjusted analysis ^94^ |
| Longest quit (years) | General population UK |  | - The odds of cessation at 4 weeks were not significantly different as longest quit (years, continuous) increased, in analyses adjusted for all other variables including the Consideration of Future Consequences (CFCS) scale (OR 1.07, 95% CI 0.99 to 1.16), CFCS future subscale (OR 1.07, 95% CI 0.99 to 1.16) and CFCS immediate subscale (OR 1.07, 95% CI 0.99 to 1.16) ^68^ | - The odds of cessation at 6 months were significantly higher as longest quit (years, continuous) increased, in analyses adjusted for all other variables including the Consideration of Future Consequences (CFCS) scale (OR 1.14, 95% CI 1.05 to 1.24), CFCS future subscale (OR 1.14, 95% CI 1.05 to 1.24) and CFCS immediate subscale (OR 1.14, 95% CI 1.05 to 1.24) ^68^ |
|  | Mental health UK Canada | No data | No data | No data |
|  | Secondary care UK Canada | No data | No data | No data |
| Time since start of most recent quit attempt | General population UK |  | - The odds of cessation at 6 months were not significantly different for those whose most recent quit attempt was between a week and a month ago (OR 1.03, 95% CI 0.67 to 1.59), 1-2 months ago (OR 0.96, 95% CI 0.62 to 1.48), and 2-3 months ago (OR 1.07, 95% CI 0.70 to 1.66) than for those whose previous quit attempt was the previous week, in an unadjusted analysis, and for those whose most recent quit attempt was between a week and a month ago (OR 0.80, 95% CI 0.51 to 1.28), 1-2 months ago (OR 0.68, 95% CI 0.43 to 1.08), 2-3 months ago (OR 0.68, 95% CI 0.43 to 1.09), and 3-6 months ago (OR 0.93, 95% CI 0.60 to 1.44) than for those whose most recent quit attempt was the previous week, in an adjusted analysis ^74^ | - The odds of cessation at 6 months were significantly higher for those whose most recent quit attempt was 3-6 months ago than for those whose most recent quit attempt was the previous week, in an unadjusted analysis (OR 1.68, 95% CI 1.14 to 2.52) ^74^ |
|  | Mental health UK Canada | No data | No data | No data |
|  | Secondary care UK Canada | No data | No data | No data |
| **Intervention characteristics** | | | | |
| Spontaneous quit attempt | General population UK |  | - The odds of cessation at 6 months were not significantly different for those who did than for those who did not make a spontaneous quit attempt, in an adjusted analysis (OR 1.19, 95% CI 0.95 to 1.49) ^74^ | - The odds of cessation at 6 months were significantly higher for those who did than for those who did not make a spontaneous quit attempt, in an unadjusted analysis (OR 1.31, 95% CI 1.07 to 1.60) ^74^ |
|  | Mental health UK Canada | No data | No data | No data |
|  | Secondary care UK Canada | No data | No data | No data |
| Smoking reduction prior to quitting | General population UK |  | - The odds of cessation at 4 weeks were not significantly different for those who quit abruptly (one RCT intervention arm) than for those who reduced first (another RCT intervention arm), in analyses adjusted for all other variables including the Consideration of Future Consequences (CFCS) scale (OR 1.37, 95% CI 0.99 to 1.89), CFCS future subscale (OR 1.37, 95% CI 1.00 to 1. 09) and CFCS immediate subscale (OR 1.37, 95% CI 0.99 to 1.89) ^68^ - The odds of cessation at 6 months were not significantly different for those who quit abruptly (one RCT intervention arm) than for those who reduced first (another RCT intervention arm), in analyses adjusted for all other variables including the Consideration of Future Consequences (CFCS) scale (OR 1.41, 95% CI 0.93 to 2.15), CFCS future subscale (OR 1.42, 95% CI 0.93 to 2.15) and CFCS immediate subscale (OR 1.41, 95% CI 0.93 to 2.13) ^68^ | - The odds of cessation at 6 months were significantly higher for those who did than for those who did not cut down first, in unadjusted (OR 3.19, 95% CI 2.58 to 3.97) and adjusted (OR 3.08, 95% CI 2.46 to 3.88) analyses ^74^ - The odds of cessation at 4 weeks to 6 months were significantly higher for those who reported smoking reduction prior to quitting than for those who did not, in unadjusted (OR 1.55, 95% CI 1.19 to 2.01) and adjusted (OR 1.49, 95% CI 1.14 to 1.94) analyses ^67^ |
|  | Mental health UK Canada | No data | No data | No data |
|  | Secondary care UK Canada | No data | No data | No data |
| Smoking reduction with NRT first | General population UK |  |  | - The odds of cessation at 4 weeks to 6 months were significantly higher for those who reported smoking reduction with NRT prior to quitting than for those who did not, in unadjusted (OR 1.47, 95% CI 1.03 to 2.10) and adjusted (OR 1.51, 95% CI 1.06 to 2.16) analyses ^67^ |
|  | Mental health UK Canada | No data | No data | No data |
|  | Secondary care UK Canada | No data | No data | No data |
| Temporary abstinence with NRT first | General population UK |  |  | - The odds of cessation at 4 weeks to 6 months were significantly higher for those who reported temporary abstinence with NRT prior to quitting than for those who did not, in unadjusted (OR 1.98, 95% CI 1.44 to 2.71) and adjusted (OR 2.10, 95% CI 1.52 to 2.89) analyses ^67^ |
|  | Mental health UK Canada | No data | No data | No data |
|  | Secondary care UK Canada | No data | No data | No data |
| Received support from cessation clinic or helpline | General population UK |  |  | - The odds of cessation at 28 days were significantly higher for those who received support from a cessation clinic or helpline than no support in an adjusted analysis (OR 1.92, 95% CI 1.20 to 3.06) ^75^ |
|  | Mental health UK Canada | No data | No data | No data |
|  | Secondary care UK Canada | No data | No data | No data |
| Received help from a doctor or other health professional | General population UK |  | - The odds of cessation at 28 days were not significantly different for those who received doctor or other health professional, than no support in an adjusted analysis (OR 0.72, 95% CI 0.43 to 1.19) ^75^ |  |
|  | Mental health UK Canada | No data | No data | No data |
|  | Secondary care UK Canada | No data | No data | No data |
| Received pharmacotherapy plus help from a clinic, doctor, or other HP | General population UK |  |  | - The odds of cessation at 28 days were significantly higher for those who received pharmacotherapy plus help from a clinic, doctor, or other health professional, than no support in an adjusted analysis (OR 2.13, 95% CI 1.58 to 2.87) ^75^ |
|  | Mental health UK Canada | No data | No data | No data |
|  | Secondary care UK Canada | No data | No data | No data |
| Face-to-face behavioural support (as a cessation aid) | General population UK |  | - The odds of cessation from the start of the most recent quit attempt to the time of the survey were not significantly different for those who did than for those who did not have face-to-face behavioural cessation support in an analysis adjusted for smoking cessation aid variables but no covariates (OR 1.02, 95% CI 0.84 to 1.24), an analysis adjusted for all covariates but no other smoking cessation aid variables (OR 1.24, 95% CI 1.00 to 1.55) and a fully adjusted analysis (OR 1.20, 95% CI 0.95 to 1.50) ^79^ |  |
|  | Mental health UK Canada | No data | No data | No data |
|  | Secondary care UK Canada | No data | No data | No data |
| Telephone support (as a cessation aid) | General population UK | - The odds of cessation from the start of the most recent quit attempt to the time of the survey were significantly lower for those who did than for those who did not have telephone cessation support in a fully adjusted analysis, in a subsample of those in social grades C2DE (OR 0.40, 95% CI 0.16 to 0.99) ^79^ | - The odds of cessation from the start of the most recent quit attempt to the time of the survey were not significantly different for those who did than for those who did not have telephone cessation support in an analysis adjusted for smoking cessation aid variables but no covariates (OR 0.66, 95% CI 0.39 to 1.12), including in subsamples of those with low and high cigarette addition and social grades ABC1 and C2DE; an analysis adjusted for all covariates but no other smoking cessation aid variables (OR 0.83, 95% CI 0.47 to 1.47) including in subsamples of those with low and high cigarette addition and social grades ABC1 and C2DE; and a fully adjusted analysis (OR 0.75, 95% CI 0.42 to 1.35) including in subsamples of those with low and high cigarette addition and social grades ABC1 ^79^ |  |
|  | Mental health UK Canada | No data | No data | No data |
|  | Secondary care UK Canada | No data | No data | No data |
| Written self-help materials (as a cessation aid) | General population UK |  | - The odds of cessation from the start of the most recent quit attempt to the time of the survey were not significantly different for those who did than for those who did not have written self-help materials (as a cessation aid) in an analysis adjusted for smoking cessation aid variables but no covariates (OR 0.91, 95% CI 0.66 to 1.26), including in subsamples of those with low and high cigarette addition; an analysis adjusted for all covariates but no other smoking cessation aid variables (OR 0.92, 95% CI 0.64 to 1.32) including in subsamples of those with low and high cigarette addition; and a fully adjusted analysis (OR 0.91, 95% CI 0.63 to 1.32) including in subsamples of those with low and high cigarette addition ^79^ |  |
|  | Mental health UK Canada | No data | No data | No data |
|  | Secondary care UK Canada | No data | No data | No data |
| Websites (as a cessation aid) | General population UK |  | - The odds of cessation from the start of the most recent quit attempt to the time of the survey were not significantly different for those who did than for those who did not have websites (as a cessation aid) in an analysis adjusted for smoking cessation aid variables but no covariates (OR 1.07, 95% CI 0.73 to 1.57), including in subsamples of those aged 16-44 and ≥45 years, those with low and high cigarette addition and those in social grades ABC1 and C2DE; an analysis adjusted for all covariates but no other smoking cessation aid variables (OR 1.19, 95% CI 0.78 to 1.83) including in subsamples of those aged 16-44 and ≥45 years, those with low and high cigarette addition and those in social grades ABC1; and a fully adjusted analysis (OR 1.25, 95% CI 0.81 to 1.92) including in subsamples of those aged 16-44 and ≥45 years, those with low and high cigarette addition and those in social grades ABC1 ^79^ | - The odds of cessation from the start of the most recent quit attempt to the time of the survey were significantly higher for those who did than for those who did not have websites (as a cessation aid), in a subsample of those in social grades C2DE in an analysis adjusted for all covariates but no other smoking cessation aid variables (OR 2.14, 95% CI 1.18 to 3.87) and a fully adjusted analysis (OR 2.20, 95% CI 1.22 to 3.98) (Jackson 2019)^79^ |
|  | Mental health UK Canada | No data | No data | No data |
|  | Secondary care UK Canada | No data | No data | No data |
| SC support setting | General population UK | - The odds of cessation at 4 weeks were significantly lower for those who received cessation support in a GP practice (OR 0.58, 95% CI 0.52 to 0.64), in a pharmacy (OR 0.89, 95% CI 0.80 to 0.98), and in a Stop Smoking Service (OR 0.71, 95% CI 0.57 to 0.89) than in the community in an adjusted analysis ^87^ - The odds of cessation at 4 weeks were significantly lower for those who received cessation support in a primary care setting than in specialist clinics (Stop Smoking Services) (OR 0.83, 95% CI 0.75 to 0.91) ^70^ - The odds of cessation at 4 weeks were significantly lower for those who received cessation support in a primary care setting than in specialist clinics (OR 0.80, 95% CI 0.66 to 0.99) ^69^ | - The odds of cessation at 4 weeks were not significantly different for those who received cessation support in an ‘other’ setting (not pharmacy, GP practice, Stop Smoking Service or the community) than those who received cessation support in the community (OR 0.88, 95% CI 0.77 to 1.02) in an adjusted analysis ^87^ - The odds of cessation at 4 weeks were not significantly different for those who received cessation support in an ‘other’ setting (not primary care or specialist clinic) than in specialist clinics (Stop Smoking Services) (OR 1.03, 95% CI 0.94 to 1.13) in an adjusted analysis ^70^ - The odds of cessation at 4 weeks were not significantly different for those who received cessation support in in a primary care setting (OR 1.02, 95% CI 0.81 to 1.27) and an ‘other’ setting (not primary care or specialist clinic; OR 0.99, 95% CI 0.77 to 1.27) than in specialist clinics (Stop Smoking Services) in an adjusted analysis of a sub-sample including only those with HSI data ^70^ - The odds of cessation at 4 weeks were not significantly different for those who received cessation support in specialist clinics than for those who received cessation support in another setting (OR 1.01, 95% CI 0.99 to 1.04) in an adjusted analysis ^71^ - The odds of cessation at 4 weeks were not significantly different for those who received cessation support in in a pharmacy setting (OR 0.94, 95% CI 0.83 to 1.06) and an ‘other’ setting (not primary care, pharmacy or specialist clinic; OR 0.87, 95% CI 0.90 to 1.08) than in specialist clinics in an adjusted analysis ^69^ | - The odds of cessation at 4 weeks were significantly higher for those receiving one-to-one cessation support in the community than for those receiving group cessation support in a specialist clinic in univariate analysis (OR 1.2, p=0.031), adjusted analysis including all variables reaching significance in the univariate analyses (OR 1.36, p=0.012) and an adjusted analysis including all variables reaching significance in the univariate analyses alongside choice of NRT product (OR 1.46, p=0.002) ^85^ |
|  | Mental health UK Canada | No data | No data | No data |
|  | Secondary care UK Canada | No data | No data | No data |
| Group support | General population UK |  |  | - The odds of cessation at 4 weeks were significantly higher for those with group support than other support in an adjusted model (OR 1.39, 95% CI 1.32 to 1.46) ^71^ - The odds of cessation at 4 weeks were significantly higher for those in a closed group (OR 1.43, 95% CI 1.16 to 1.76) and those in open (rolling) groups (OR 1.46, 95% CI 1.19 to 1.78) than for those receiving one-to-one support in an adjusted analysis ^69^ |
|  | Mental health UK Canada |  | - The odds of cessation at end of treatment were not significantly different for those at least one session of group therapy than for those receiving no group therapy (OR 2.43, 95% CI 0.94 to 6.25) in an adjusted analysis of the whole sample, including those with and without schizophrenia ^91^ |  |
|  | Secondary care UK Canada | No data | No data | No data |
| One-to-one cessation support | General population UK |  | - The odds of cessation at 4 weeks were not significantly different for those receiving other support than for those receiving one-to-one support (OR 0.97, 95% CI 0.68 to 1.38) in an unadjusted analysis ^72^ |  |
|  | Mental health UK Canada | No data | No data | No data |
|  | Secondary care UK Canada | No data | No data | No data |
| Drop-in cessation support | General population UK | - The odds of cessation at 4 weeks were significantly lower for those receiving drop-in cessation support (OR 0.72, 95% CI 0.57 to 0.90) than for those receiving one-to-one support in an adjusted analysis ^69^ |  |  |
|  | Mental health UK Canada | No data | No data | No data |
|  | Secondary care UK Canada | No data | No data | No data |
| Number of contacts / treatment sessions | General population UK |  | - The odds of cessation at 1 year were not significantly different for those who had an unknown number of contacts at the smoking cessation clinic (OR 1.6, 95% CI 0.6 to 4.6) than for those who had 0-3 contacts, in an adjusted model of the English smoking cessation services ^77^ | - The odds of cessation at 1 year were significantly higher for those who had 4-6 contacts (OR 2.0, 95% CI 1.3 to 3.2) and 7-15 contacts at the smoking cessation clinic (OR 2.6, 95% CI 1.4 to 4.7) than for those who had 0-3 contacts, in an adjusted model of the English smoking cessation services ^77^ |
|  | Mental health UK Canada |  | - The odds of cessation at 12 to 26 weeks were not significantly different for those with a total greater number of visits to the programme (continuous) (OR 1.0, 95% CI 0.9 to 1.1) in a univariate analysis ^88^ | - The odds of cessation at the end of treatment were significantly higher among those who had received 13-24 (OR 3.95, 95% CI 1.14 to 13.64) and ≥25 treatment sessions (OR 4.12, 95% CI 1.23 to 13.77) than for those who had received 1-12 individual treatment sessions ^91^ - The odds of cessation at 8 to 28 weeks were significantly higher as the total number of treatment visits (continuous) increased in an unadjusted analysis (OR 1.07, 95% CI 1.04 to 1.10), an adjusted analysis of the whole sample (OR 1.08, 95% CI 1.04 to 1.12), an adjusted analysis of the substance use disorder subsample (OR 1.08, 95% CI 1.01 to 1.15), an adjusted analysis of a psychiatric disorder sub-sample (OR 1.13, 95% CI 1.02 to 1.25), and an adjusted analysis of the sub-sample of those with co-occurring substance use and psychiatric disorders (OR 1.07, 95% CI 1.04 to 1.11) ^90^ - The odds of cessation at 8 to 26 weeks were significantly higher as the total number of visits to the programme (continuous) increased in an adjusted analysis of the whole sample (OR 1.17, 95% CI 1.12 to 1.23), an adjusted analysis of a female subsample (OR 1.13, 95% CI 1.06 to 1.22), and an adjusted analysis of a male sub-sample (OR 1.20, 95% CI 1.12 to 1.28) ^89^ |
|  | Secondary care UK Canada |  | - The odds of cessation at most recent follow-up were not significantly different for those with a greater number of total visits to the programme (continuous) in an unadjusted analysis (OR 1.1, 95% CI 1.0 to 1.3) ^94^ |  |
| Length of time in the programme | General population UK | No data | No data | No data |
|  | Mental health UK Canada | No data | No data | No data |
|  | Secondary care UK Canada |  | - The odds of cessation at most recent follow-up were not significantly different for those with a greater length of time in the programme (continuous) in unadjusted (OR 1.0, 95% CI 1.0 to 1.1) or adjusted (OR 1.0, 95% CI 1.0 to 1.1) analyses ^94^ |  |
| Practitioner - number of treatment episodes delivered | General population UK |  | - The odds of cessation at 4 weeks was not significantly different for those seeing practitioners who had delivered 101-200, 201-300, 301-400, 401-500 or >500 treatment episodes than for those seeing practitioners who has delivered 1-100 treatment episodes, in both an adjusted analysis of the whole sample, and an adjusted analysis of a sub-sample that had HSI data ^70^ |  |
|  | Mental health UK Canada | No data | No data | No data |
|  | Secondary care UK Canada | No data | No data | No data |
| Received pharmacotherapy | General population UK |  | - The odds of cessation at 6 months were not significantly different for those who received medication on prescription combined with brief advice than for those who received none of the listed cessation aids in unadjusted (OR 1.20, 95% CI 0.89 to 1.62) and partially adjusted (OR 1.38, 95% CI 1.00 to 1.91) analyses ^84^ | - The odds of cessation at 28 days were significantly higher for those who received pharmacotherapy than no support in an adjusted analysis (OR 1.50, 95% CI 1.18 to 1.90) ^75^ - The odds of cessation at 6 months were significantly higher for those who received medication on prescription combined with behavioural support than for those who received none of the listed cessation aids in unadjusted (OR 1.98, 95% CI 1.20 to 3.24), partially adjusted (OR 2.27, 95% CI 1.32 to 3.92) and fully adjusted (OR 2.58, 95% CI 1.48 to 4.52) analyses, and for those who received medication on prescription combined with brief advice than for none of the cessation aids only in the fully adjusted analysis (OR 1.55, 95% CI 1.11 to 2.16) ^84^ |
|  | Mental health UK Canada |  | - The odds of cessation at 12 to 26 weeks were not significantly different for those who received combination therapy (any combination of NRT or oral medications during treatment) than those who received a single product or no pharmacotherapy in a univariate analysis (OR 1.50, 95% CI 0.2 to 14.0) ^88^ - The odds of cessation at 8 to 28 weeks were not significantly different for those who received combination therapy than those who received monotherapy in an unadjusted analysis (OR 0.85, 95% CI 0.59 to 1.22), nor in an adjusted analysis of the subsample consisting of those with a psychiatric disorder (OR 0.30, 95% CI 0.08 to 1.17) ^90^ - The odds of cessation at the end of treatment were not significantly different with combination therapy than with monotherapy or no pharmacotherapy in an adjusted analysis of the whole sample, including those with and without schizophrenia (OR 2.06, 95% CI 0.73 to 5.81) ^91^ |  |
|  | Secondary care UK Canada | No data | No data | No data |
| Prescription NRT (as a cessation aid) | General population UK | - The odds of cessation from the start of the most recent quit attempt to the time of the survey were significantly lower for those with prescription NRT than for those with no NRT among those aged 16-44 years in an analysis adjusted for smoking cessation aid variables but no covariates (OR 0.96, 95% CI 0.55 to 0.87) ^79^ | - The odds of cessation from the start of the most recent quit attempt to the time of the survey were not significantly different for those with prescription NRT than for those with no NRT (OR 0.88, 95% CI 0.76 to 1.02) and for those with prescription NRT than for those with no NRT among those aged ≥45 years (OR 1.04, 95% CI 0.85 to 1.28) in an analysis adjusted for smoking cessation aid variables but no covariates, nor for those with prescription NRT than for those with no NRT among those aged 16-44 years in an analysis adjusted for all covariates but no other smoking cessation aid variables (OR 1.05, 95% CI 0.82 to 1.36) and a fully adjusted analysis adjusted (OR 1.09, 95% CI 0.85 to 1.42) ^79^ | - The odds of cessation from the start of the most recent quit attempt to the time of the survey were significantly higher for those with prescription NRT than for those with no NRT in an analysis adjusted for all covariates but no other smoking cessation aid variables (OR 1.28, 95% CI 1.08 to 1.52) and a fully adjusted analysis adjusted (OR 1.34, 95% CI 1.12 to 1.59), and for those with prescription NRT than for those with no NRT among those aged ≥45 years in an analysis adjusted for all covariates but no other smoking cessation aid variables (OR 1.50, 95% CI 1.19 to 1.88) and a fully adjusted analysis adjusted (OR 1.58, 95% CI 1.25 to 2.00) ^79^ - The odds of cessation at 4 weeks were significantly higher for those who had combined than single NRTs with behavioural support in an adjusted analysis (OR 1.82, 95% CI 1.07 to 3.09) ^82^ - The odds of cessation at 4 weeks were significantly higher for those who had single NRT (OR 1.39, 95% CI 1.25 to 1.54) and combination NRT (OR 2.82, 95% CI 2.56 to 3.12) than for those who had no medication, and for those who had combination NRT (OR 2.45, 95% CI 1.83 to 3.28) than for those who had no medication in an adjusted analysis of a subsample of only those with HSI data ^70^ |
|  | Mental health UK Canada | No data | No data | No data |
|  | Secondary care UK Canada | No data | No data | No data |
| NRT bought over the counter (as a cessation aid) | General population UK | - The odds of cessation from the start of the most recent quit attempt to the time of the survey were significantly lower for those who had NRT bought over the counter than for those who had no NRT in an analysis adjusted for smoking cessation aid variables but no covariates (OR 0.68, 95% CI 0.61 to 0.75) and an analysis adjusted for all covariates but no other smoking cessation aid variables (OR 0.88, 95% CI 0.79 to 0.99) ^79^ - The odds of cessation at 6 months were significantly lower for those who received NRT bought over the counter than for those who received none of the listed cessation aids in unadjusted (OR 0.57, 95% CI 0.42 to 0.78), partially adjusted (OR 0.62, 95% CI 0.45 to 0.85) and fully adjusted (OR 0.68, 95% CI 0.49 to 0.94) analyses ^84^ | - The odds of cessation from the start of the most recent quit attempt to the time of the survey were not significantly different for those who had NRT bought over the counter than for those who had no NRT in a fully adjusted analysis (OR 0.98, 95% CI 0.87 to 1.09) ^79^ |  |
|  | Mental health UK Canada | No data | No data | No data |
|  | Secondary care UK Canada | No data | No data | No data |
| NRT use (no details on whether prescribed or bought) | General population UK |  |  | - The odds of cessation at 4 weeks were significantly higher for those who received single NRT than those who received no medication in an adjusted analysis (OR 1.75, 95% CI 1.39 to 2.22) ^69^ |
|  | Mental health UK Canada |  | - The odds of cessation at 8 to 28 weeks were not significantly different as number of weeks of patch use (continuous; OR 1.02, 95% CI 1.00 to 1.05), number of weeks of gum use (continuous; OR 1.01, 95% CI 0.98 to 1.04), number of weeks of inhaler use (continuous; OR 1.01, 95% CI 0.97 to 1.05) and number of weeks of lozenge use (continuous; OR 1.03, 95% CI 0.98 to 1.09) increased in an unadjusted analysis, nor as number of weeks of patch use (continuous) increased in an adjusted analysis (OR 0.99, 95% CI 0.96 to 1.02) ^90^ |  |
|  | Secondary care UK Canada | No data | No data | No data |
| Varenicline (as a cessation aid) | General population UK |  |  | - The odds of cessation from the start of the most recent quit attempt to the time of the survey were significantly higher for those who did than for those who did not have varenicline in an analysis adjusted for smoking cessation aid variables but no covariates (OR 1.31, 95% CI 1.11 to 1.54), an analysis adjusted for all covariates but no other smoking cessation aid variables (OR 1.67, 95% CI 1.38 to 2.01) and a fully adjusted analysis (OR 1.82, 95% CI 1.51 to 2.21) ^79^ - The odds of cessation at 4 weeks were significantly higher for those who had varenicline than for those who had NRT (OR 1.82, 95% CI 1.61 to 2.06) in an adjusted analysis ^87^ - The odds of cessation at 4 weeks were significantly higher for those who had varenicline than for those who had no medication in an adjusted analysis of the whole sample (OR 3.09, 95% CI 2.79 to 3.41) and an adjusted analysis of a subsample of only those with HSI data (OR 2.73, 95% CI 2.03 to 3.68) ^70^ - The odds of cessation at 4 weeks were significantly higher for those who received varenicline (OR 1.78, 95% CI 1.57 to 2.02) and combination NRT (OR 1.42, 95% CI 1.06 to 1.91) than those who received single NRT in an adjusted analysis ^69^ |
|  | Mental health UK Canada |  | - The odds of cessation at 8 to 28 weeks were not significantly different as number of weeks of varenicline use (continuous) increased (OR 1.02, 95% CI 0.99 to 1.06) in an unadjusted analysis ^90^ |  |
|  | Secondary care UK Canada | No data | No data | No data |
| Bupropion (as a cessation aid) | General population UK |  | - The odds of cessation from the start of the most recent quit attempt to the time of the survey were not significantly different for those who did than for those who did not have bupropion in an analysis adjusted for smoking cessation aid variables but no covariates (OR 0.74, 95% CI 0.52 to 1.05), an analysis adjusted for all covariates but no other smoking cessation aid variables (OR 1.23, 95% CI 0.83 to 1.81) and a fully adjusted analysis (OR 1.27, 95% CI 0.86 to 1.89) ^79^ - The odds of cessation at 4 weeks were not significantly different for those who received bupropion than those who received single NRT in an adjusted analysis (OR 1.128, 95% CI 0.96 to 1.30) ^69^ | - The odds of cessation at 4 weeks were significantly higher for those who had bupropion than for those who had no medication in an adjusted analysis of the whole sample (OR 2.29, 95% CI 1.82 to 2.89) and an adjusted analysis of a subsample of only those with HSI data (OR 2.12, 95% CI 1.31 to 3.46) ^70^ - The odds of cessation at 4 weeks were significantly higher for those who chose bupropion as a cessation aid than those who chose NRT in univariate analysis (OR 1.39, p=0.005) and in an adjusted analysis including all variables reaching significance in the univariate analyses (OR 1.40, p=0.012) ^85^ |
|  | Mental health UK Canada |  | - The odds of cessation at 8 to 28 weeks were not significantly different as number of weeks of bupropion use (continuous) increased (OR 0.78, 95% CI 0.52 to 1.17) in an unadjusted analysis ^90^ |  |
|  | Secondary care UK Canada | No data | No data |  |
| E-cigarettes (as a cessation aid) | General population UK |  | - The odds of cessation between 6 and 12 months were not significantly different for those who had e-cigarettes than those who had over-the-counter NRT in an analysis adjusted for age, sex, social grade, time since quit attempt started, quit attempts in the past year, abrupt versus gradual quitting, year of the survey, time spent with urges to smoke and strength of urges to smoke (Model 3; OR 1.64, 95% CI 0.83 to 3.24) ^72^ - The odds of cessation at up to 1 year were not significantly different for those who had e-cigarettes than those who had no cessation aid in an analysis adjusted for age, sex, social grade, time since quit attempt started, quit attempts in the past year, abrupt versus gradual quitting and year of the survey (Model 2; OR 1.21, 95% CI 0.92 to 1.58) ^72^ - The odds of cessation between 6 and 12 months were not significantly different for those who had e-cigarettes than those who had no cessation aid in an unadjusted analysis (Model 1; OR 1.18, 95% CI 0.72 to 1.94), an analysis adjusted for age, sex, social grade, time since quit attempt started, quit attempts in the past year, abrupt versus gradual quitting and year of the survey (Model 2; OR 0.91, 95% CI 0.54 to 1.55), and an analysis adjusted for Model 2 variables plus time spent with urges to smoke and strength of urges to smoke (Model 3; OR 1.10, 95% CI 0.59 to 2.06) ^72^ | - The odds of cessation from the start of the most recent quit attempt to the time of the survey were significantly higher for those who did than for those who did not have e-cigarettes in an analysis adjusted for smoking cessation aid variables but no covariates (OR 1.49, 95% CI 1.34 to 1.67), including in a sub-sample of men (OR 1.76, 95% CI 1.51 to 2.04) and women (OR 1.25, 95% CI 1.06 to 1.47); an analysis adjusted for all covariates but no other smoking cessation aid variables (OR 1.86, 95% CI 1.62 to 2.13), including in a sub-sample of men (OR 2.15, 95% CI 1.78 to 2.60) and women (OR 1.58, 95% CI 1.29 to 1.94); and a fully adjusted analysis (OR 1.95, 95% CI 1.69 to 2.24) ), including in a sub-sample of men (OR 2.26, 95% CI 1.87 to 2.74) and women (OR 1.66, 95% CI 1.65 to 2.04) ^79^ - The odds of cessation at up to 1 year were significantly greater for those who had e-cigarettes than those who had over-the-counter NRT in an unadjusted analysis (Model 1; OR 2.23, 95% CI 1.70 to 2.93), an analysis adjusted for age, sex, social grade, time since quit attempt started, quit attempts in the past year, abrupt versus gradual quitting and year of the survey (Model 2; OR 1.88, 95% CI 1.40 to 2.52), an analysis adjusted for Model 2 variables plus time spent with urges to smoke and strength of urges to smoke (Model 3; OR 1.63, 95% CI 1.17 to 2.28), and an analysis adjusted for Model 3 variables plus the interaction terms time since last quit attempt started x time spent with urges and time since last quit attempt started x strength of urges to smoke (Model 4; OR 1.63, 95% CI 1.17 to 2.27) ^72^ - The odds of cessation at up to 6 months were significantly greater for those who had e-cigarettes than those who had over-the-counter NRT in an unadjusted analysis (Model 1; OR 2.06, 95% CI 1.50 to 2.82), an analysis adjusted for age, sex, social grade, time since quit attempt started, quit attempts in the past year, abrupt versus gradual quitting and year of the survey (Model 2; OR 1.80, 95% CI 1.27 to 2.55), and an analysis adjusted for Model 2 variables plus time spent with urges to smoke and strength of urges to smoke (Model 3; OR 1.56, 95% CI 1.06 to 2.29) ^72^ - The odds of cessation between 6 and 12 months were significantly greater for those who had e-cigarettes than those who had over-the-counter NRT in an unadjusted analysis (Model 1; OR 2.56, 95% CI 1.49 to 4.42), and an analysis adjusted for age, sex, social grade, time since quit attempt started, quit attempts in the past year, abrupt versus gradual quitting and year of the survey (Model 2; OR 1.98, 95% CI 1.11 to 3.53 (Brown 2014)^72^ - The odds of cessation at up to 1 year were significantly greater for those who had e-cigarettes than those who had no cessation aid in an unadjusted analysis (Model 1; OR 1.38, 95% CI 1.08 to 1.76), an analysis adjusted for age, sex, social grade, time since quit attempt started, quit attempts in the past year, abrupt versus gradual quitting, year of the survey, time spent with urges to smoke and strength of urges to smoke (Model 3; OR 1.62, 95% CI 1.19 to 2.19), and an analysis adjusted for Model 3 variables plus the interaction terms time since last quit attempt started x time spent with urges and time since last quit attempt started x strength of urges to smoke (Model 4; OR 1.61, 95% CI 1.19 to 2.18) ^72^ - The odds of cessation at up to 6 months were significantly greater for those who had e-cigarettes than those who had no cessation aid in an unadjusted analysis (Model 1; OR 1.49, 95% CI 1.12 to 1.98), an analysis adjusted for age, sex, social grade, time since quit attempt started, quit attempts in the past year, abrupt versus gradual quitting and year of the survey (Model 2; OR 1.39, 95% CI 1.01 to 1.90), and an analysis adjusted for Model 2 variables plus time spent with urges to smoke and strength of urges to smoke (Model 3; OR 1.88, 95% CI 1.32 to 2.68) ^72^ |
|  | Mental health UK Canada | No data | No data | No data |
|  | Secondary care UK Canada | No data | No data | No data |
| Any e-cigarette use at baseline | General population UK |  | - The odds of cessation at 1 year were not significantly different for those with e-cigarette use than no e-cigarette use at baseline (OR 0.83, 95% CI 0.52 to 1.30) ^78^ |  |
|  | Mental health UK Canada | No data | No data | No data |
|  | Secondary care UK Canada | No data | No data | No data |
| E-cigarette type and frequency of use at follow-up | General population UK | - The odds of cessation at 1 year were lower for those who used a non-daily cigalike than those with no e-cigarette use (OR 0.35, 95% CI 0.20 to 0.60) in an adjusted analysis ^78^ | - The odds of cessation at 1 year were not significantly different for those who used a daily cigalike (OR 0.74, 95% CI 0.39 to 1.42) and non-daily tank (OR 0.70, 95% CI 0.29 to 1.68) than for those with no e-cigarette use in an adjusted analysis ^78^ | - The odds of cessation at 1 year were significantly higher for those who used a daily tank (OR 2.69, 95% CI 1.48 to 4.89) than for those with no e-cigarette use in an adjusted analysis ^78^ |
|  | Mental health UK Canada | No data | No data | No data |
|  | Secondary care UK Canada | No data | No data | No data |
| Compliance with pharmacotherapy | General population UK |  | - The odds of cessation at 1 year were not significantly different for those for whom there was no evidence of compliance (OR 2.3, 95% CI 1.0 to 4.9) than for those who were non-compliant (i.e. <4 weeks) with pharmacotherapy in an adjusted model ^77^ | - The odds of cessation at 1 year were significantly higher for those who did not receive pharmacotherapy (OR 4.9, 95% CI 1.5 to 15.5), were compliant with pharmacotherapy for 4-6 weeks (OR 3.0, 95% CI 1.4 to 6.1), and were compliant with pharmacotherapy for ≥7 weeks (OR 4.8, 95% CI 2.3 to 10.3), than for those who were non-compliant (i.e. <4 weeks) with pharmacotherapy in an adjusted model ^77^ - The odds of cessation at 1 year were significantly higher for those who finished NRT or sessions than for those who did not finish either (OR 9.5, 95% CI 3.0 to 30.2) in an adjusted analysis ^77^ |
|  | Mental health UK Canada | No data | No data | No data |
|  | Secondary care UK Canada | No data | No data | No data |
| Treatment x setting interaction term | General population UK |  | - The odds of cessation at 4 weeks were not significantly different according to the interaction terms NRT x community, NRT x GP practice, NRT x pharmacy, NRT x Stop Smoking Service, and NRT x other setting (all OR 1.0, 95% CI not reported) in an adjusted analysis ^87^ - The odds of cessation at 12 weeks were not significantly different according to the interaction terms varenicline x community (OR 1.0, 95% CI not reported), varenicline x GP practice (OR 0.93, 95% CI 0.77 to 1.13), varenicline x Stop Smoking Service (OR 0.78, 95% CI 0.62 to 2.08), and varenicline x other setting (OR 1.15, 95% CI 0.79 to 1.58) in an adjusted analysis ^87^ |  |
|  | Mental health UK Canada | No data | No data | No data |
|  | Secondary care UK Canada | No data | No data | No data |
| Treatment x FTND interaction term | General population UK |  | - The odds of cessation at 4 weeks were not significantly different according to the interaction terms NRT x FTND (OR 1.0, 95% CI not reported), and varenicline x FTND (OR 0.97, 95% CI 0.94 to 1.01) in an adjusted analysis ^87^ |  |
|  | Mental health UK Canada | No data | No data | No data |
|  | Secondary care UK Canada | No data | No data | No data |
